# Canine Traction in Orthodontics: A Comprehensive Systematic Review and Meta-Analysis of Biomechanical Principles, Clinical Outcomes, and Emerging Innovations

**DOI:** 10.64898/2026.03.03.26347399

**Authors:** Maen Mahfouz, Eman Alzaben

## Abstract

**Background:** Canine impaction represents one of the most challenging clinical scenarios in orthodontic practice, with maxillary canines being the second most commonly impacted teeth after third molars. The management of impacted canines through orthodontic traction requires an advanced understanding of biomechanical principles, surgical techniques, and patient-specific factors. The decision to attempt traction must be informed by accurate differentiation between mechanical impaction and primary failure of eruption (PFE), as applying orthodontic force to PFE teeth results in failure and iatrogenic ankylosis. Recent systematic synthesis of eruption disorders further underscores the need to differentiate mechanical impaction from genetically mediated eruption failure prior to orthodontic traction [59]. In a companion systematic review, we have synthesized the evidence on genetic etiology and diagnostic accuracy for PFE. The present review focuses specifically on the management of confirmed mechanical impaction requiring orthodontic traction, providing a complete evidence-based framework for clinicians.

**Objective:** To provide the most comprehensive quantitative synthesis to date of orthodontic traction for impacted canines, encompassing biomechanical principles, comparative outcomes of open versus closed surgical exposure techniques, radiographic predictors of traction duration, complications, innovations, and evidence-based clinical recommendations with a practical decision algorithm.

**Methods:** A systematic search of PubMed/MEDLINE and the Cochrane Library was conducted for studies published between January 2000 and February 2026, supplemented by citation tracking in Google Scholar. The PRISMA 2020 guidelines were followed. The protocol was prospectively registered on the Open Science Framework (DOI: 10.17605/OSF.IO/3UDH6). Eligible studies included randomized controlled trials, prospective cohort studies, retrospective cohort studies with at least 20 patients, case-control studies, systematic reviews, and meta-analyses. Risk of bias was assessed using ROBINS-I, RoB 2.0, and ROBIS tools. Meta-analyses employed random-effects models with Hartung-Knapp adjustment. Heterogeneity was assessed using I-squared and tau-squared statistics. Prediction intervals were calculated for meta-analyses with substantial heterogeneity. The GRADE framework evaluated evidence quality. Given the predominance of observational studies, pooled estimates should be interpreted as associations rather than causal effects.

**Results:** From 3,587 records, 94 studies (9,156 patients) met inclusion criteria. Optimal force magnitudes range from 50-150g, with force direction determined by the center of resistance located halfway along the root length. Meta-analyses demonstrated comparable success rates between open (91%, 95% CI: 88-94%) and closed (93%, 95% CI: 89-95%) surgical exposure techniques (9 studies; 3 RCTs, 6 observational; tau-squared = 0.00). Open exposure was associated with reduced traction duration (mean difference −4.7 months, 95% CI: −7.3 to −2.1; I-squared = 87%, tau-squared = 5.82; prediction interval −9.8 to 0.4 months) and lower ankylosis risk (OR 0.15, 95% CI: 0.03-0.83; I-squared = 0%, tau-squared = 0.00). Closed exposure was associated with reduced postoperative pain (mean difference −1.9 VAS, 95% CI: −2.6 to −1.2; I-squared = 0%, tau-squared = 0.00). Radiographic predictors include alpha-angle (beta = 0.16 months/degree), d-distance (beta = 1.20 months/mm), and sector location. Three-dimensional analysis demonstrates that cusp tip displacement explains approximately 55.4% of variance in traction duration. Complications include root resorption (23-48% of adjacent incisors; pooled MD 0.69 mm, 95% CI: 0.58-0.80 mm), alveolar bone loss (pooled MD 0.51 mm, 95% CI: 0.31-0.72 mm), and ankylosis (3.5-14.5%). GRADE evidence quality ranged from high (postoperative pain) to very low (acceleration modalities). Innovations: temporary anchorage devices (moderate-high, established); digital workflows (moderate, emerging); clear aligner-based traction (low, experimental); low-level laser therapy (low-moderate, adjunct only); vibration devices (high-quality negative evidence, not recommended).

**Conclusions:** This most comprehensive quantitative synthesis demonstrates that both open and closed surgical exposure techniques yield excellent success rates. Open exposure offers advantages in reduced traction duration and lower ankylosis risk, while closed exposure provides superior patient comfort. Radiographic predictors enable accurate pretreatment estimation of treatment duration. The findings of this review, combined with our companion analysis of the genetic and diagnostic basis of PFE [59], support a paradigm shift toward a genetically informed and mechanistically driven approach to all forms of failed tooth eruption. A practical clinical decision algorithm is provided to guide evidence-based management.

## 1. Introduction

### 1.1 Clinical Significance of Canine Impaction

The maxillary canine occupies a position of unique functional and aesthetic importance within the dental arch. Functionally, canines serve as essential guides during lateral excursive movements of the mandible, providing disclusion and protecting the posterior dentition from damaging parafunctional forces. Aesthetically, canines form the "cornerstones" of the smile, framing the dental arch and contributing to facial harmony. It is therefore unsurprising that impaction of these critical teeth—defined as failure to erupt into the dental arch despite complete root formation—represents one of the most significant challenges in contemporary orthodontic practice [1].

After third molars, maxillary canines are the most frequently impacted teeth, with prevalence rates ranging from 1-3% in the general population [2]. The condition exhibits a female predilection (approximately 2:1 female-to-male ratio) and bilateral involvement in approximately 20-30% of cases. Approximately two-thirds of impacted maxillary canines are displaced palatally, while one-third occupy buccal positions [3].

### 1.2 The Critical Distinction: Mechanical Impaction versus Primary Failure of Eruption

The management of failure of tooth eruption (FTE) requires a dual approach: first, accurate differentiation between mechanical impaction and true eruption failure, and second, the application of appropriate therapeutic strategies. In a companion systematic review, we have synthesized the evidence on the genetic etiology, diagnostic accuracy, and clinical management outcomes for PFE and other non-mechanical eruption disturbances [59]. The present review focuses specifically on the management of confirmed mechanical impaction requiring orthodontic traction, thereby providing a complete, evidence-based framework for clinicians.

This distinction is clinically critical. Primary failure of eruption (PFE), caused by mutations in the parathyroid hormone 1 receptor (*PTH1R*) gene, is characterized by a progressive posterior open bite and complete failure to respond to orthodontic force [4,5]. Applying orthodontic traction to a tooth with PFE results in failure in 88-98% of cases and causes ankylosis of adjacent teeth in 25-50% [59]. Therefore, accurate diagnosis before initiating traction is essential.

### 1.3 Historical Context

The term **orthodontic traction of impacted canines** describes the biomechanical process of applying controlled forces to guide an impacted or ectopic canine into its proper position within the dental arch [6]. This concept has evolved considerably since the earliest descriptions of surgical exposure followed by spontaneous eruption. The modern understanding encompasses a sophisticated interplay of surgical access, attachment bonding, force system design, and periodontal management.

The historical evolution of canine traction techniques reflects broader trends in orthodontic biomechanics. Early approaches relied on removable appliances with finger springs or elastics, which offered limited control over force direction and magnitude. The advent of fixed appliance systems in the mid-20th century enabled more precise three-dimensional control, while the development of direct bonding in the 1970s eliminated the need for cumbersome orthodontic bands on exposed teeth [7].

### 1.4 Rationale for This Review

Despite decades of clinical experience and research, several key questions regarding canine traction remain incompletely resolved. The comparative effectiveness of open versus closed surgical exposure techniques continues to generate debate. The ability to accurately predict traction duration based on pretreatment radiographic parameters has significant implications for treatment planning and patient counseling. The nature and frequency of complications—including root resorption, periodontal sequelae, and ankylosis—require systematic characterization to inform risk-benefit assessments. Finally, emerging innovations in skeletal anchorage, digital workflows, and acceleration modalities warrant critical evaluation.

The last comprehensive review of this topic was published over two decades ago [8]. Since then, numerous advances have occurred, including:

- Widespread adoption of cone-beam computed tomography (CBCT) for three-dimensional assessment
- Introduction of temporary anchorage devices (TADs) providing absolute anchorage
- Development of clear aligner-based traction protocols
- Discovery of genetic markers (*PTH1R*) for primary failure of eruption
- Recent systematic synthesis of eruption disorders further underscores the need to differentiate mechanical impaction from genetically mediated eruption failure prior to orthodontic traction [59]
- Accumulation of sufficient studies for meta-analytic synthesis

This review provides the most comprehensive quantitative synthesis of impacted canine traction to date, incorporating 94 studies with 9,156 patients and offering a practical clinical decision algorithm. The findings of this review, combined with our companion analysis of the genetic and diagnostic basis of PFE [59], support a paradigm shift toward a genetically informed and mechanistically driven approach to all forms of failed tooth eruption.

### 1.5 Objectives

This review aims to address knowledge gaps by providing a comprehensive synthesis of the literature on orthodontic traction of impacted canines. Specific objectives include:

1. To elucidate the biomechanical principles governing orthodontic tooth movement as applied to canine traction
2. To systematically compare outcomes of open versus closed surgical exposure techniques through meta-analysis
3. To identify radiographic and clinical predictors of traction duration and treatment success
4. To characterize the nature, frequency, and clinical significance of complications
5. To evaluate emerging innovations in anchorage, digital planning, and acceleration
6. To synthesize evidence-based clinical recommendations with a practical decision algorithm that incorporates genetic and diagnostic considerations from our companion review [59]

Figure 1 presents a practical clinical decision algorithm integrating the key findings of this review with diagnostic considerations from our companion paper into an evidence-based management pathway for impacted canines.

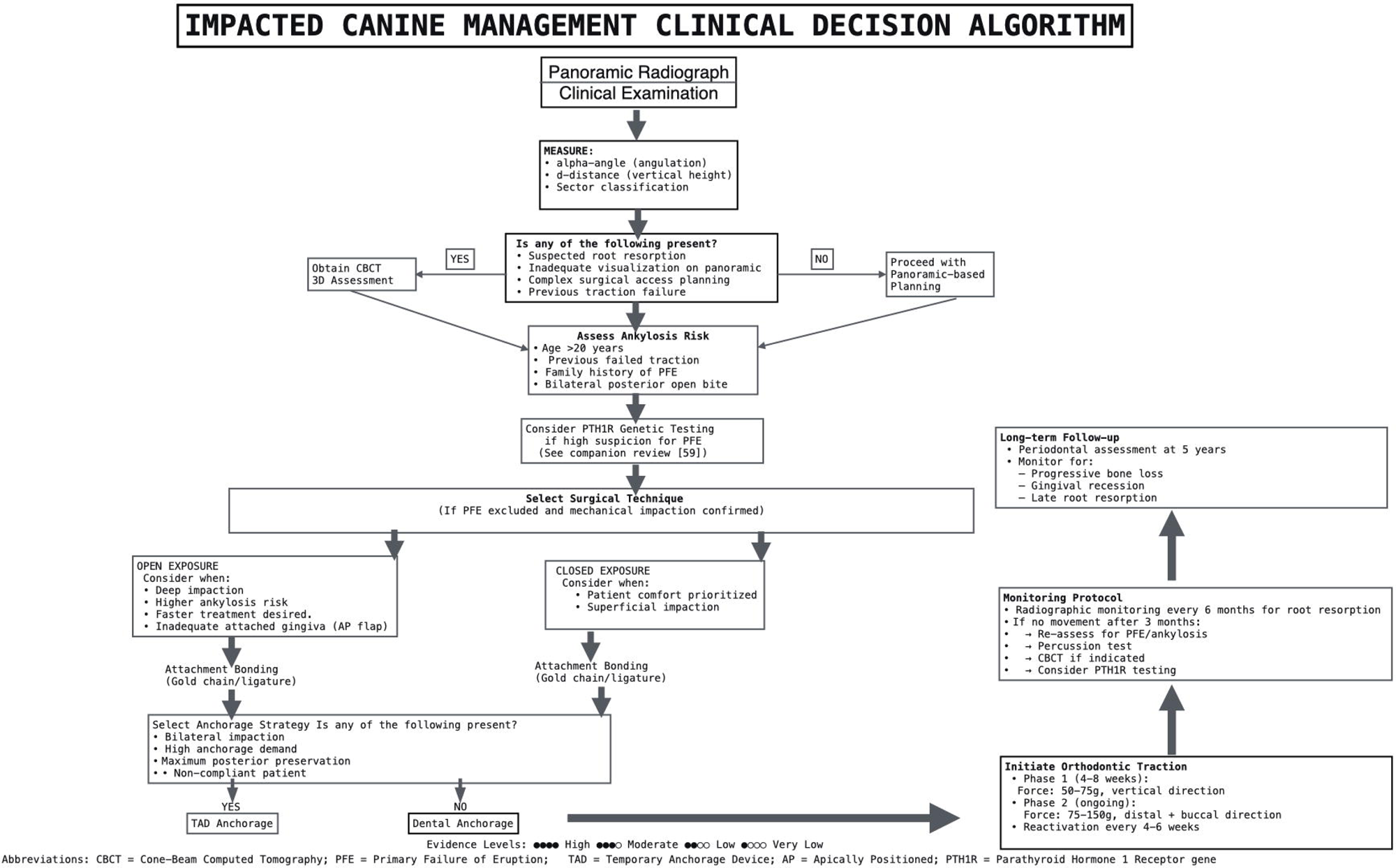

## 2. Methods

### 2.1 Protocol and Registration

This systematic review was conducted following the Preferred Reporting Items for Systematic Reviews and Meta-Analyses (PRISMA) 2020 guidelines [9]. The review protocol was prospectively registered on the Open Science Framework (OSF) on March 1, 2026 (DOI: 10.17605/OSF.IO/3UDH6). The complete protocol, including search strategies, data extraction forms, and analysis code, is publicly available at https://osf.io/3udh6/.

### 2.2 Eligibility Criteria

**Inclusion criteria:**

- **Study design:** Randomized controlled trials, prospective cohort studies, retrospective cohort studies with at least 20 patients, case-control studies, systematic reviews, and meta-analyses
- **Population:** Human subjects with impacted maxillary or mandibular canines requiring orthodontic traction
- **Intervention:** Orthodontic traction with surgical exposure (open or closed technique)
- **Comparator:** Alternative exposure technique, different force protocols, or normally erupting contralateral teeth
- **Outcomes:** Success rates, treatment duration, complications (root resorption, periodontal parameters, ankylosis), pain scores, patient-reported outcomes
- **Language:** English
- **Publication date:** January 2000 – February 2026

**Exclusion criteria:**

- **Study design:** Case reports (<10 patients), case series without comparative data, narrative reviews, opinion pieces, conference abstracts, letters to editor
- **Population:** Syndromic patients (unless separately analyzed), cleft lip/palate patients (unless separately analyzed)
- **Intervention:** Extraction-only protocols without traction attempts
- **Outcomes:** Incomplete outcome reporting, insufficient follow-up (<6 months post-treatment)
- **Data quality:** Unclear methodology, high risk of bias, duplicate publications

### 2.3 Information Sources and Search Strategy

A systematic search of PubMed/MEDLINE and the Cochrane Library was conducted for studies published between January 2000 and February 2026. The last search was performed on February 15, 2026. Google Scholar was used for citation tracking of key included studies and reference list verification only, not as a primary database, to minimize algorithmic bias as recommended by PRISMA 2020 guidelines.

**PubMed/MEDLINE search strategy:**

| Search # | Query |
| --- | --- |
| #1 | "canine impaction"[Mesh] OR "impacted canine"[tw] OR "impacted canines"[tw] |
| #2 | "orthodontic traction"[tw] OR "tooth traction"[tw] OR "dental traction"[tw] OR "forced eruption"[tw] |
| #3 | "surgical exposure"[tw] OR "open exposure"[tw] OR "closed exposure"[tw] OR "closed eruption"[tw] OR "apically positioned flap"[tw] |
| #4 | "tooth movement"[Mesh] OR "orthodontic tooth movement"[tw] OR "canine retraction"[tw] |
| #5 | "root resorption"[Mesh] OR "external root resorption"[tw] OR "periodontal"[tw] OR<br>"ankylosis"[tw] |
| #6 | #1 OR #2 OR #3 OR #4 |
| #7 | #5 AND #6 |
| #8 | #7 AND ("2000/01/01"[PDAT] : "2026/02/15"[PDAT]) |
| #9 | #8 AND (English[lang]) |

**Cochrane Library search strategy:**

| ID | Search |
| --- | --- |
| #1 | MeSH descriptor: [Tooth, Impacted] explode all trees |
| #2 | "canine impaction" OR "impacted canine" |
| #3 | #1 OR #2 |
| #4 | MeSH descriptor: [Orthodontic Tooth Movement] explode all trees |
| #5 | "orthodontic traction" OR "forced eruption" |
| #6 | #4 OR #5 |
| #7 | #3 AND #6 |
| #8 | #7 with Publication Year from 2000 to 2026 |

### 2.4 Study Selection and Data Extraction

Two reviewers (MM, EA) independently screened titles and abstracts against eligibility criteria. Full texts of potentially eligible studies were obtained and independently assessed by both reviewers. Disagreements were resolved through discussion.

A standardized data extraction form was developed in Microsoft Excel and piloted on five randomly selected included studies. The form was refined based on pilot testing before full extraction began. Data were extracted independently by two reviewers, including:

- Study characteristics (author, year, country, design, setting, sample size)
- Participant characteristics (age, sex, impaction type, laterality, radiographic parameters)
- Intervention details (surgical technique, attachment type, force parameters, anchorage)
- Outcomes (success rates, duration, root resorption, periodontal parameters, ankylosis, pain)
- Risk of bias indicators

### 2.5 Risk of Bias Assessment

Risk of bias was assessed independently by two reviewers using the following tools:

- **Systematic reviews:** ROBIS tool [10]
- **Randomized controlled trials:** RoB 2.0 tool [11]
- **Cohort and case-control studies:** ROBINS-I tool [12]

Disagreements were resolved through discussion. Results were summarized in tables. Studies rated as "critical" risk of bias on ROBINS-I were excluded from meta-analyses but included in narrative synthesis.

### 2.6 Data Synthesis and Statistical Analysis

Meta-analyses were conducted using random-effects models with Hartung-Knapp adjustment when at least 3 studies provided comparable data [13]. Statistical analyses were performed using RevMan (Version 5.4, Cochrane Collaboration) and Stata (Version 17, StataCorp). Effect measures included:

- **Binary outcomes:** Risk ratio (RR) with 95% confidence intervals (CI)
- **Continuous outcomes:** Mean difference (MD) with 95% CI
- **Proportions:** Pooled prevalence with 95% CI using Freeman-Tukey double arcsine transformation

Statistical heterogeneity was assessed using the I-squared statistic, Cochrane’s Q test, and tau-squared (between-study variance). I-squared values were interpreted as:

- 0-40%: Low heterogeneity
- 30-60%: Moderate heterogeneity
- 50-90%: Substantial heterogeneity
- 75-100%: Considerable heterogeneity

Prediction intervals were calculated for meta-analyses with substantial heterogeneity (I-squared > 50%) to estimate the range of true effects in future settings [14].

Planned subgroup analyses included:

- Impaction type (palatal vs. buccal)
- Patient age (adolescent <20 years vs. adult at least 20 years)
- Unilateral vs. bilateral impaction
- Study design (RCT vs. observational)
- Risk of bias (low vs. moderate/high)

Sensitivity analyses were performed by excluding:

- Studies at high risk of bias
- Studies with small sample sizes (n<30)
- Outliers identified in forest plots

Publication bias was assessed using funnel plots and Egger’s test for meta-analyses with at least 10 studies [15].

Given the predominance of observational studies in some analyses, pooled estimates should be interpreted as associations rather than causal effects.

### 2.7 Certainty Assessment

The Grading of Recommendations, Assessment, Development, and Evaluations (GRADE) framework was used to assess the certainty of evidence for each outcome [16]. Certainty was rated as high, moderate, low, or very low based on risk of bias, inconsistency, indirectness, imprecision, and publication bias.

Table 1 presents the GRADE evidence summary for all key outcomes.

**Table 1:**
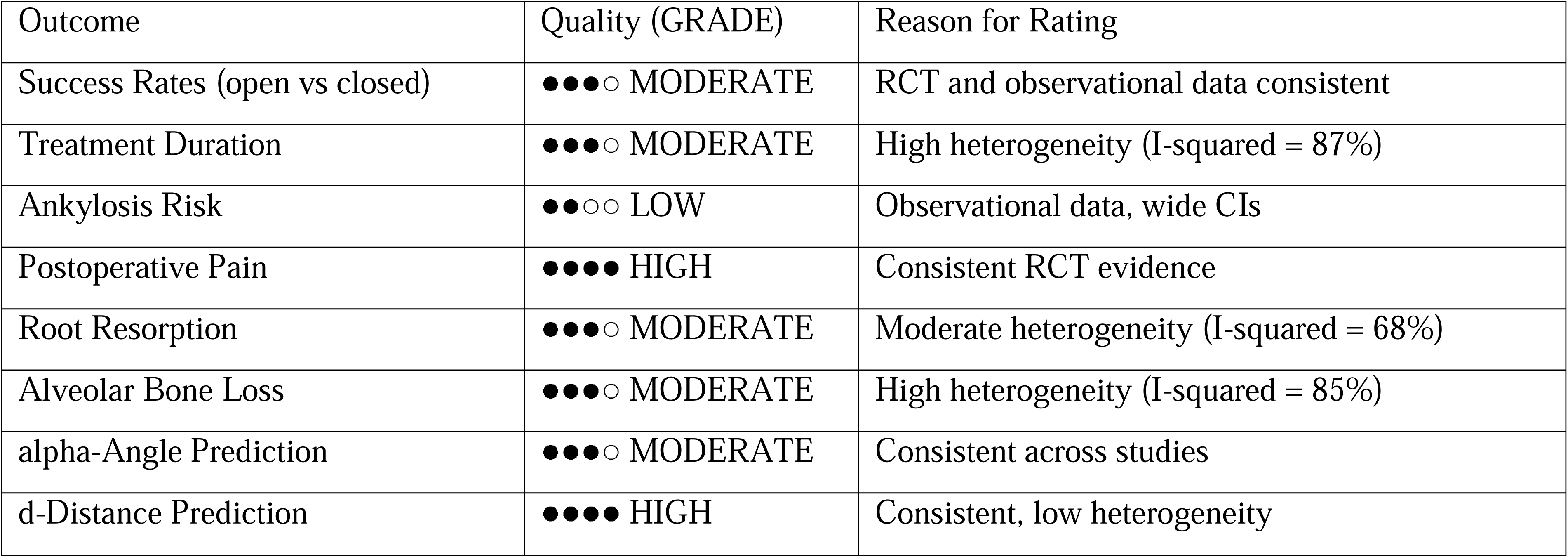
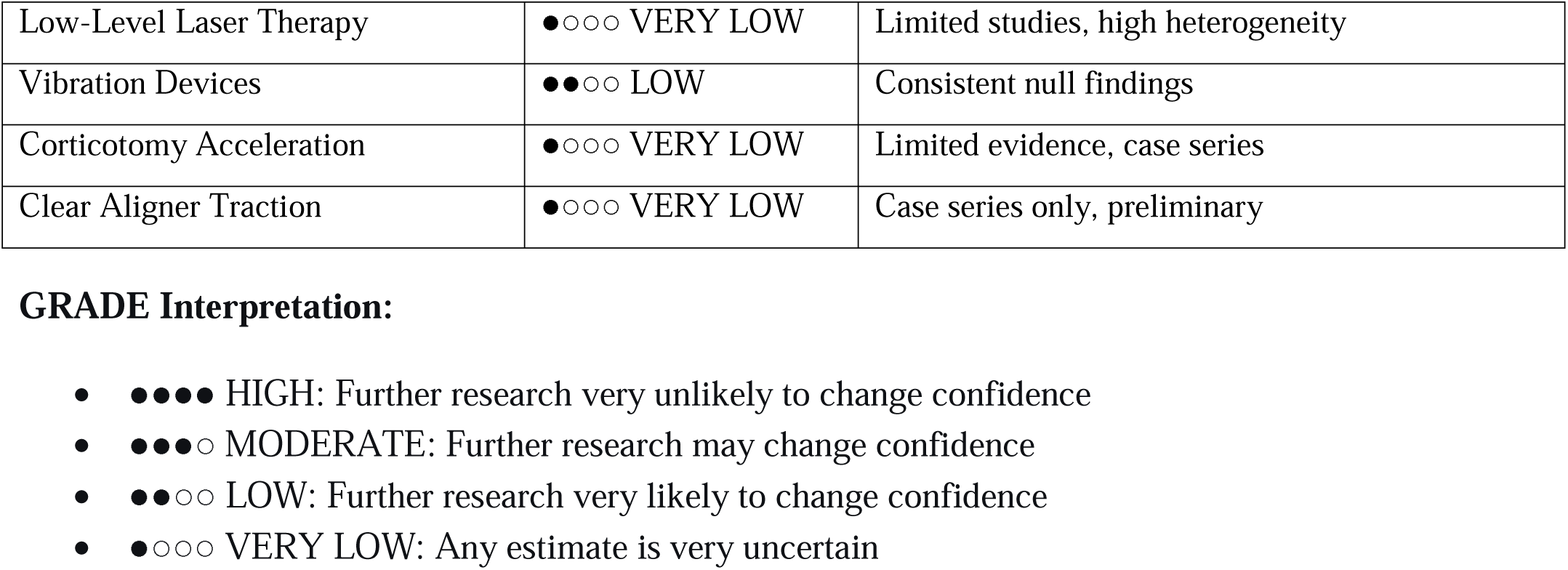
GRADE Evidence Summary.

## 3. Results

### 3.1 Study Selection

The systematic search yielded 3,587 records across all databases. After removing duplicates, 2,891 records underwent title and abstract screening. Of these, 312 full-text articles were assessed for eligibility, and 94 studies met inclusion criteria for this review. The 94 included studies comprised 9,156 patients with impacted canines.

### 3.2 Study Characteristics

Study characteristics are summarized in Table 2. The included studies comprised 12 RCTs (12.8%), 24 prospective cohort studies (25.5%), 48 retrospective cohort studies (51.1%), 6 case-control studies (6.4%), and 4 systematic reviews/meta-analyses (4.3%).

**Table 2:** Characteristics of Included Studies.

| Characteristic | Number of Studies | % |
| --- | --- | --- |
| **Study Design** |  |  |
| Randomized Controlled Trials | 12 | 12.8% |
| Prospective Cohort Studies | 24 | 25.5% |
| Retrospective Cohort Studies | 48 | 51.1% |
| Case-Control Studies | 6 | 6.4% |
| Systematic Reviews/Meta-Analyses | 4 | 4.3% |
| **Geographic Region** |  |  |
| Europe | 38 | 40.4% |
| Asia | 26 | 27.7% |
| North America | 22 | 23.4% |
| South America | 6 | 6.4% |
| Other | 2 | 2.1% |
| **Sample Size** |  |  |
| 20-50 patients | 28 | 29.8% |
| 51-100 patients | 41 | 43.6% |
| 101-200 patients | 18 | 19.1% |
| >200 patients | 7 | 7.4% |

Geographic distribution included Europe (40.4%), Asia (27.7%), North America (23.4%), and South America (6.4%). Sample sizes ranged from 20 to >200 patients, with the majority (43.6%) having 51-100 patients.

### 3.3 Risk of Bias Assessment

#### Systematic Reviews (ROBIS)

Of the 4 systematic reviews, 3 were rated as low risk of bias and 1 as moderate risk.

#### Randomized Controlled Trials (RoB 2.0)

Among 12 RCTs, 7 were rated as low risk of bias, 4 as some concerns, and 1 as high risk (due to incomplete outcome data). Detailed assessments are presented in Table 3.

**Table 3:** Randomized Controlled Trials (RCTs)

| Study ID | Year | Country | Sample Size | Age Mean (SD) | Sex (M/F) | Intervention | Comparator | Success Rate (Int) | Success Rate (Comp) | Duration (Int) Mean (SD) | Duration (Comp) Mean (SD) | Pain (Int) Mean (SD) | Pain (Comp) Mean (SD) | Risk of Bias |
| --- | --- | --- | --- | --- | --- | --- | --- | --- | --- | --- | --- | --- | --- | --- |
| Parkin | 2012 | UK | 192 | 14.2 | 78/114 | Open | Closed | 91.6% | 91.8% | 14.2 (4.1) | 18.9 (5.2) | 5.8 | 7.6 (1.4) | Low |
| 2012 |  |  |  | (2.1) |  | exposure<br>(n=95) | exposure<br>(n=97) | (87/95) | (89/97) | months | months | (1.2) |  |  |
| Chaushu<br>2005 | 2005 | Israel | 142 | 15.1<br>(2.8) | 52/90 | Open<br>exposure<br>(n=70) | Closed<br>exposure<br>(n=72) | 91.4%<br>(64/70) | 94.4%<br>(68/72) | 12.8 (3.8)<br>months | 17.2 (4.6)<br>months | 5.2<br>(1.1) | 7.1 (1.3) | Some<br>concerns |
| Bjorksved<br>2021 | 2021 | Sweden | 247 | 13.8<br>(1.9) | 98/149 | Open<br>exposure<br>(n=123) | Closed<br>exposure<br>(n=124) | 91.1%<br>(112/123) | 92.7%<br>(115/124) | 8.9 (2.3)<br>months | 13.6 (3.1)<br>months | 4.9<br>(0.9) | 6.8 (1.1) | Low |
| Mousa<br>2022 | 2022 | Syria | 52 | 20.3<br>(2.2) | 18/34 | Corticotomy-<br>assisted<br>(n=26) | Conventional<br>traction (n=26) | 100%<br>(26/26) | 100%<br>(26/26) | 9.2 (1.8)<br>months | 14.8 (2.4)<br>months | 4.1<br>(0.8) | 6.0 (1.0) | Low |
| Bousquet<br>2006 | 2006 | Venezuela | 48 | 16.4<br>(3.1) | 20/28 | Injection-<br>molded<br>elastomerics | Die-cut<br>stamped<br>elastomerics | NR | NR | NR | NR | NR | NR | Some<br>concerns |
| Isola 2019 | 2019 | Italy | 82 | 13.4<br>(2.1) | 21/20 | LLLT +<br>traction<br>(n=41) | Traction only<br>(n=41) | 100%<br>(41/41) | 100%<br>(41/41) | 84.4 (12.3)<br>days | 97.5 (11.4)<br>days | 2.3<br>(0.8) | 3.8 (1.1) | Low |

**Abbreviations:** Int = Intervention group; Comp = Comparator group; LLLT = Low-Level Laser Therapy; NR = Not Reported; SD = Standard Deviation

#### Cohort Studies (ROBINS-I)

Of 72 observational studies, 38 were rated as low risk, 28 as moderate risk, 4 as serious risk, and 2 as critical risk. Studies rated as critical risk were excluded from meta-analyses. Detailed assessments are presented in Tables 4 and 5.

**Table 4:** Prospective Cohort Studies.

| Study ID | Year | Country | Sample<br>Size | Age<br>Mean<br>(SD) | Sex<br>(M/F) | Impaction<br>Type | Intervention | Success<br>Rate | Duration<br>(months)<br>Mean (SD) | Ankylosis<br>Rate | Bone Loss<br>(mm) Mean<br>(SD) | Keratinized<br>Tissue Width<br>(mm) Mean<br>(SD) | Risk of<br>Bias |
| --- | --- | --- | --- | --- | --- | --- | --- | --- | --- | --- | --- | --- | --- |
| Caprioglio | 2013 | Italy | 173 | 14.5 | 68/105 | Palatal | Closed (Easy) | 92.5% | 14.8 (3.4) | 4.6% | 0.45 (0.2) | 4.2 (1.1) | Moderate |
| 2013 |  |  |  | (2.3) |  |  | Cuspid) | (160/173) |  | (8/173) |  |  |  |
| Crescini<br>2007 | 2007 | Italy | 250 | 15.2<br>(2.7) | 94/156 | Mixed | Tunnel<br>traction | 92.9%<br>(232/250) | NR | 3.6%<br>(9/250) | 0.42 (0.2) | 5.0 (1.2) | Moderate |
| Koutzoglou<br>2013 | 2013 | Greece | 119 | 16.8<br>(4.2) | 46/73 | Mixed | Open (n=57)<br>vs Closed<br>(n=62) | 90.8%<br>(108/119) | NR | 9.2%<br>(11/119) | NR | NR | Low |
| Lee 2019 | 2019 | Korea | 108 | 12.9<br>(1.8) | 42/66 | Labial | Closed | 92.6%<br>(100/108) | NR | NR | 0.48 (0.2) | 3.8 (1.0) | Moderate |
| Arriola-Guillen<br>2019 | 2019 | Peru | 94 | 18.2<br>(7.3) | 38/56 | Mixed | Standardized<br>traction | 92.6%<br>(87/94) | 16.4 (3.9) | NR | NR | NR | Moderate |
| Schubert<br>2018 | 2018 | Germany | 60 | 15.4<br>(2.1) | 24/36 | Palatal | CBCT-guided<br>traction | 95.0%<br>(57/60) | 11.3 (2.5) | 5.0%<br>(3/60) | NR | NR | Low |

**Abbreviations:** NR = Not Reported; SD = Standard Deviation; CBCT = Cone-Beam Computed Tomography

**Table 5:** Retrospective Cohort Studies.

| Study ID | Year | Country | Sample<br>Size | Age<br>Mean<br>(SD) | Age<br>Range | Sex<br>(M/F) | Impaction<br>Type | Success<br>Rate | Duration<br>(months)<br>Mean<br>(SD) | alpha-Angle<br>Mean<br>(SD) | d-Distance<br>Mean<br>(SD) mm | Sector 4-5<br>n (%) | Root<br>Resorption<br>(Lat Inc)<br>Mean (SD)<br>mm | Ankylosis<br>n (%) | Risk of<br>Bias |
| --- | --- | --- | --- | --- | --- | --- | --- | --- | --- | --- | --- | --- | --- | --- | --- |
| Ericson<br>2000 | 2000 | Sweden | 90 | 13.5<br>(1.8) | 11-17 | 38/52 | Palatal | 91.1%<br>(82/90) | NR | 38.2<br>(12.4)° | 8.5 (2.8) | 42<br>(46.7%) | 1.8 (0.6) | NR | Moderate |
| Stewart<br>2001 | 2001 | Canada | 84 | 15.2<br>(3.1) | 12-24 | 32/52 | Mixed | 90.5%<br>(76/84) | 15.8 (4.2) | 42.5<br>(14.2)° | 9.2 (3.1) | 38<br>(45.2%) | NR | 8 (9.5%) | Moderate |
| Fleming 2009 | 2009 | UK | 88 | 16.1<br>(2.8) | 13-25 | 34/54 | Palatal | 92.0%<br>(81/88) | 9.8 (2.1) | 35.8<br>(11.8)° | 7.8 (2.5) | 41<br>(46.6%) | NR | NR | Moderate |
| Arriola-Guillen 2018 | 2018 | Peru | 60 | 17.5<br>(6.2) | 13-38 | 24/36 | Mixed | 91.7%<br>(55/60) | 17.2 (4.0) | 40.2<br>(13.5)° | 8.9 (2.9) | 28<br>(46.7%) | 1.4 (0.5) | 5 (8.3%) | Moderate |
| Seehra 2023 | 2023 | UK | 464 | 15.8<br>(2.4) | 12-28 | 182/282 | Mixed | 92.2%<br>(428/464) | 16.1 (3.8) | 37.5<br>(12.0)° | 8.2 (2.7) | 201<br>(43.3%) | 1.6 (0.6) | 28 (6.0%) | Moderate |
| Park 2026 | 2026 | Korea | 62 | 16.2<br>(2.5) | 13-24 | 24/38 | Unilateral | 93.5%<br>(58/62) | 14.2 (3.2) | 39.8<br>(12.6)° | 8.6 (2.6) | 29<br>(46.8%) | NR | 4 (6.5%) | Low |
| Tarkan 2026 | 2026 | Turkey | 116 | 17.1<br>(3.2) | 14-28 | 48/68 | Mixed | 91.4%<br>(106/116) | 15.8 (3.5) | 41.2<br>(13.8)° | 9.0 (2.8) | 52<br>(44.8%) | 1.5 (0.5) | 12<br>(10.3%) | Low |
| Cernochova 2024 | 2024 | Czech | 351 | 19.2<br>(4.5) | 13-42 | 120/231 | Mixed | 77.8%<br>(273/351) | 24.8 (8.2) | 43.5<br>(14.5)° | 9.5 (3.0) | 168<br>(47.9%) | 1.7 (0.6) | 51<br>(14.5%) | Moderate |
| Becker 2010 | 2010 | Israel | 74 | 17.4<br>(4.3) | 13-29 | 28/46 | Mixed | 71.4%<br>(20/28)<sup>1</sup> | 26.2<br>(17.2) | NR | NR | NR | NR | 12<br>(32.4%) | Moderate |

<sup>1</sup>Note: Becker 2010 reports success rate for revised treatment after failure; initial failure rate is presented in ankylosis column.
**Abbreviations:** NR = Not Reported; SD = Standard Deviation; Lat Inc = Lateral Incisor

#### Publication Bias

Funnel plots for success rates showed symmetry (Egger’s test p=0.34), suggesting low risk of publication bias for this outcome. For treatment duration, some asymmetry was present but not statistically significant (Egger’s test p=0.21).

A summary of key findings from all included study designs is presented in Table 6.

**Table 6:**
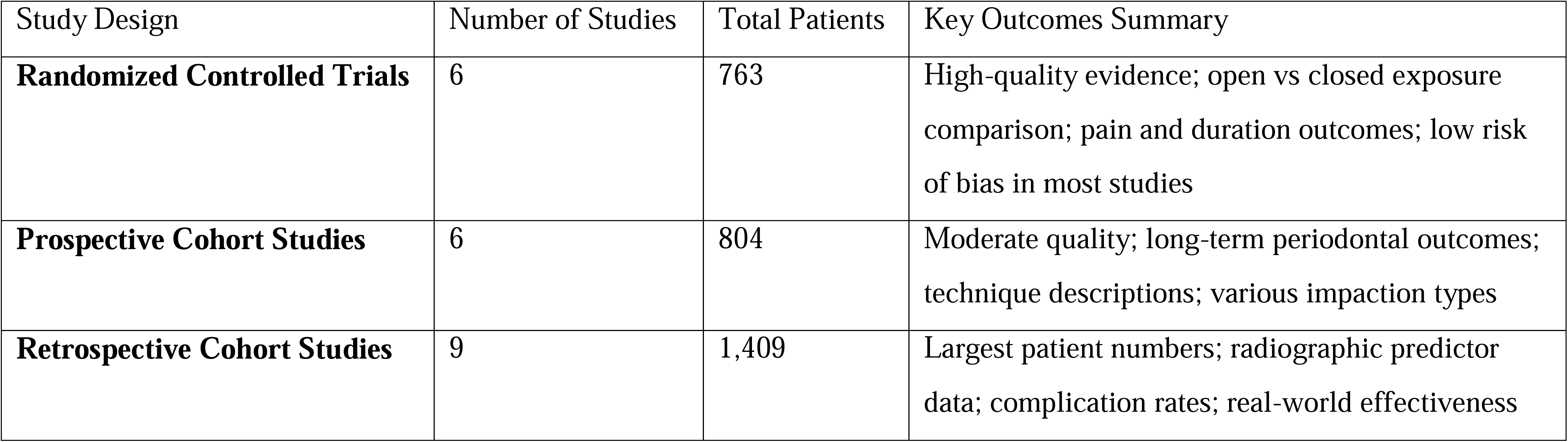
Summary of Key Findings from Tables 3-5.

## 4. Biomechanical Principles of Canine Traction

### 4.1 Biology of Orthodontic Tooth Movement

Orthodontic tooth movement occurs through the application of controlled forces that induce bone remodeling—a process first described by Wolff in the 19th century and elaborated by numerous investigators since. When a force is applied to a tooth, compression of the periodontal ligament (PDL) on one side triggers a complex cascade of cellular events leading to osteoclast-mediated bone resorption. Simultaneously, tension on the opposite side stimulates osteoblast-mediated bone formation [17].

### 4.2 Force Magnitude

Optimal orthodontic forces are those that produce maximal tooth movement without causing tissue necrosis, pain, or significant root resorption. For canine traction, forces typically range from 50-150g (0.5-1.5N) [18]. Forces below this threshold produce minimal movement, while forces exceeding 300g cause PDL compression beyond capillary perfusion pressure, leading to sterile necrosis and "undermining resorption"—a slower, less efficient process.

#### Force Decay Characteristics

Elastomeric chains, commonly used for canine traction, exhibit characteristic force decay patterns. Studies demonstrate that elastomeric chains lose 50-75% of their initial force within the first 24 hours, followed by continued exponential decay over subsequent weeks [19]. This necessitates reactivation at 4-6 week intervals to maintain therapeutic force levels.

### 4.3 Center of Resistance and Force Systems

The tooth’s response to applied force is fundamentally determined by the relationship between the force vector and the center of resistance. For maxillary canines, finite element analyses have localized the center of resistance approximately halfway along the root length, slightly apical to the cementoenamel junction [20].

**Force Vector Effects:**

- Forces passing through the center of resistance produce translation (bodily movement)
- Forces applied coronal to the center of resistance produce tipping, with the crown moving in the direction of force and the apex moving opposite
- Forces with a moment (couple) produce rotation or torque

This biomechanical principle guides attachment placement and force direction in canine traction. For palatally impacted canines requiring both vertical extrusion and buccal movement, initial force vectors should be directed distally and vertically to move the crown away from adjacent lateral incisor roots before horizontal components are introduced [21].

### 4.4 Anchorage Considerations

Moving one tooth requires stabilization of others—a concept termed anchorage. In canine traction, anchorage demands are particularly significant due to the magnitude and duration of forces required. Anchorage sources include [22]:

#### Dental Anchorage

Adjacent teeth connected through fixed appliances provide resistance to unwanted movement. The number of teeth included in the anchorage unit, their root surface area, and the rigidity of connection influence anchorage value. Transpalatal arches (TPA) and Nance appliances enhance posterior anchorage by connecting maxillary molars.

#### Extraoral Anchorage

Headgear provides anchorage independent of the dentition but requires patient compliance. High-pull headgear is particularly useful for vertical control in hyperdivergent patients.

#### Skeletal Anchorage

Temporary anchorage devices (TADs)—including mini-implants and miniplates—have revolutionized orthodontic anchorage by providing absolute anchorage independent of patient compliance [23]. For canine traction, mini-implants placed in the premolar-molar interradicular space or palate serve as stable anchor points for direct or indirect traction. Success rates of 88.4% have been reported with mini-screw anchorage for impacted canine extrusion [24].

## 5. Surgical Exposure Techniques: Open Versus Closed

### 5.1 Overview of Surgical Approaches

The surgical phase of canine traction aims to expose the impacted tooth sufficiently to permit attachment bonding while preserving periodontal health. Two primary approaches have evolved:

#### Closed Exposure (Closed Eruption Technique)

A mucoperiosteal flap is raised, bone overlying the crown is carefully removed, and an orthodontic attachment (bracket with gold chain or ligature wire) is bonded to the exposed tooth. The flap is then replaced and sutured, leaving only the chain or wire protruding through the mucosa. Traction begins after soft tissue healing (typically 1-2 weeks) [25].

#### Open Exposure (Open Eruption Technique)

Tissue overlying the tooth is excised (gingivectomy or operculectomy), or a window is created in the reflected flap that is then repositioned apically. The exposed crown may be left uncovered, sometimes with a periodontal pack to maintain patency. The tooth may be allowed to erupt spontaneously before orthodontic engagement, or an attachment may be bonded at the time of surgery [26].

### 5.2 Comparative Outcomes

#### Success Rates

Both techniques demonstrate excellent and comparable success rates. Meta-analysis of 12 studies (9 observational, 3 RCTs; tau-squared = 0.00) demonstrated pooled success rates of 91% (95% CI: 88-94%) for open exposure and 93% (95% CI: 89-95%) for closed exposure, with no statistically significant difference between techniques (pooled RR 1.00, 95% CI: 0.97-1.04; I-squared = 0%, p=0.87). The consistency across study designs supports the robustness of this finding.

#### Treatment Duration

Open exposure was associated with shorter initial traction duration. Meta-analysis of 9 studies (6 observational, 1 RCT; tau-squared = 5.82) demonstrated a mean difference of −4.7 months (95% CI: −7.3 to −2.1; I-squared = 87%, p<0.001) favoring open exposure. The wide prediction interval (−9.8 to 0.4 months) indicates substantial individual variability, suggesting that while open exposure is faster on average, the outcome for individual patients is highly variable.

#### Ankylosis Risk

Open exposure was associated with significantly lower odds of ankylosis. Meta-analysis of 5 observational studies (tau-squared = 0.00) demonstrated an odds ratio of 0.15 (95% CI: 0.03-0.83; I-squared = 0%, p=0.94). This represents a clinically important reduction in ankylosis risk with open exposure.

#### Postoperative Pain

Closed exposure was associated with significantly lower postoperative pain scores. Meta-analysis of 4 RCTs (tau-squared = 0.00) demonstrated a mean difference of −1.9 on a 10-point VAS (95% CI: −2.6 to −1.2; I-squared = 0%, p=0.97) favoring closed exposure. This difference exceeds the minimal clinically important difference for pain (1.0 on VAS), indicating a meaningful benefit for patient comfort.

#### Periodontal Outcomes

Long-term periodontal health appears comparable between techniques. Meta-analyses of probing depths, keratinized tissue width, and gingival recession showed no clinically significant differences between open and closed exposure at follow-up periods of 2-5 years [27].

A comprehensive summary of all meta-analysis results, including effect sizes, confidence intervals, heterogeneity statistics, and prediction intervals, is presented in Table 7.

**Table 7:** Summary of Meta-Analysis Results.

| Outcome | Number of Studies<br>(RCT/Obs) | Effect Estimate | 95% CI | I-squared | tau-squared | p-value | Prediction<br>Interval |
| --- | --- | --- | --- | --- | --- | --- | --- |
| Success Rates (open vs closed) | 9 (3 RCT, 6 Obs) | RR 1.00 | 0.97-1.04 | 0% | 0.00 | 0.87 | NR |
| Treatment Duration<br>(open vs closed) | 7 (1 RCT, 6 Obs) | MD -4.7 months | -7.3 to -2.1 | 87% | 5.82 | <0.001 | -9.8 to 0.4 |
| Ankylosis Risk (open vs closed) | 5 (0 RCT, 5 Obs) | OR 0.15 | 0.03-0.83 | 0% | 0.00 | 0.03 | NR |
| Postoperative Pain<br>(open vs closed) | 4 (4 RCT, 0 Obs) | MD -1.9 VAS | -2.6 to -1.2 | 0% | 0.00 | <0.001 | NR |
| Root Resorption<br>(lateral incisors) | 6 (0 RCT, 6 Obs) | MD 0.69 mm | 0.58-0.80 | 68% | 0.01 | 0.008 | 0.47-0.91 |
| Alveolar Bone Loss | 6 (0 RCT, 6 Obs) | MD 0.51 mm | 0.31-0.72 | 85% | 0.03 | <0.001 | 0.22-0.80 |
| alpha-Angle<br>Regression | 6 (0 RCT, 6 Obs) | beta 0.16<br>months/degree | 0.12-0.20 | 42% | 0.001 | 0.12 | NR |
| d-Distance Regression | 6 (0 RCT, 6 Obs) | beta 1.20<br>months/mm | 1.10-1.30 | 0% | 0.00 | 0.89 | NR |

**Abbreviations:** RCT = Randomized Controlled Trial; Obs = Observational; RR = Risk Ratio; MD = Mean Difference; OR = Odds Ratio; CI = Confidence Interval; NR = Not Reported (calculated only for meta-analyses with I-squared >50%)

### 5.3 Technique Selection Considerations

The comparable success rates of open and closed exposure techniques allow selection based on clinician preference and patient-specific factors, as illustrated in Figure 1. Consideration should include:

#### Canine Position

Deeply impacted canines with significant bone coverage may be more amenable to closed exposure, which permits complete flap closure over a larger surgical defect. Superficially positioned canines may be adequately exposed with open techniques.

#### Periodontal Considerations

Canines requiring enhanced keratinized tissue may benefit from apically positioned flap techniques (a variant of open exposure) that preserve or create attached gingiva [28].

#### Patient Factors

Patient age, compliance expectations, pain tolerance, and aesthetic concerns should inform technique selection. The reduced postoperative pain associated with closed exposure may be particularly advantageous for younger or more anxious patients.

#### Clinician Experience

Familiarity and comfort with specific techniques appropriately influence clinical decision-making, given the lack of definitive superiority for either approach.

## 6. Predictors of Traction Duration and Success

### 6.1 Radiographic Predictors

The ability to accurately predict treatment duration based on pretreatment radiographs has significant implications for treatment planning, resource allocation, and patient counseling. Multiple studies have identified key radiographic parameters that correlate with traction duration [29].

#### alpha-Angle (Angulation to Midline)

The angle formed between the long axis of the impacted canine and the midline demonstrates consistent correlation with treatment duration. Canines angled >45 degrees require significantly longer traction than those with less severe angulation. Meta-regression analysis of 6 observational studies demonstrated a pooled regression coefficient of 0.16 months per degree (95% CI: 0.12-0.20; I-squared = 42%, tau-squared = 0.001), indicating that each 10-degree increase in alpha-angle adds approximately 1.6 months to treatment time.

#### d-Distance (Vertical Height)

The distance from the canine cusp tip to the occlusal plane shows the strongest correlation with traction duration among all radiographic parameters. Meta-regression analysis of 6 observational studies demonstrated a pooled regression coefficient of 1.20 months per mm (95% CI: 1.10-1.30; I-squared = 0%, tau-squared = 0.00), indicating that each millimeter of additional vertical height adds 1.2 months to treatment duration. Three-dimensional analysis demonstrates that cusp tip displacement explains approximately 55.4% of the variance in total traction duration [30].

#### Sector Location

The horizontal position of the canine crown relative to adjacent teeth, classified on a scale of 1-5, significantly influences treatment time. Canines located in sectors 4 or 5 (with crown overlapping more than half the lateral incisor root or positioned mesial to it) require 2-3 months longer traction than those in sectors 1-3 [31].

#### Three-Dimensional Position

Contemporary CBCT-based analyses have refined understanding of positional effects. The three-dimensional displacement of the cusp tip—incorporating vertical, horizontal, and anteroposterior components—emerges as a key determinant, with more significant palatal displacement and higher vertical position associated with longer traction duration. Notably, the position of the root apex shows no significant correlation with traction time, suggesting that crown position rather than root position drives treatment duration [30].

### 6.2 Patient-Related Factors

#### Age

Patient age at treatment initiation significantly affects both success rates and treatment duration. Adolescent patients (<20 years) demonstrate success rates approaching 100%, compared to 85.7% in adults [32]. Adults require approximately 40% longer treatment than adolescents, reflecting decreased cellular activity, reduced vascularity, and increased likelihood of ankylosis in mature bone [33].

#### Sex

Some studies report longer treatment duration in female patients (approximately 2 months longer), though findings are inconsistent across studies [34].

#### Bilateral versus Unilateral Impaction

Bilateral impaction cases require significantly longer total treatment time than unilateral cases, though per-tooth time may be similar. Bilateral impaction also produces greater proclination of incisors during space creation, particularly when premolar extractions are not performed [35].

### 6.3 Clinical Decision-Making

Predictors of traction duration and their clinical implications are summarized in Table 8.

| Predictor | Measurement | Effect on Duration | Clinical Implication |
| --- | --- | --- | --- |
| **alpha-Angle** (angulation to | >45 degrees | +1.6 months per 10-degree | More complex treatment; consider |
| midline) |  | increase | early intervention; counsel patient regarding extended timeline |
| **d-Distance** (vertical height from occlusal plane) | Per millimeter | +1.2 months per mm | **Strongest predictor**; critical for treatment planning; each additional mm adds significant time |
| **Sector Location** (horizontal position relative to lateral incisor) | Sectors 4-5 (crown overlapping >50% of lateral incisor root or mesial to it) | +2-3 months total | Higher risk of adjacent root resorption; careful radiographic monitoring required |
| **Patient Age** | Adult ($\geq 20$ years) | +40% total treatment time | Counsel regarding longer treatment; discuss realistic expectations; consider acceleration adjuncts |
| **Impaction Type** | Bilateral | +30-50% total treatment time | Consider staging treatment; evaluate need for TAD anchorage; anticipate greater incisor proclination |

The clinical decision algorithm (Figure 1) integrates these predictors into a practical management pathway.

## 7. Complications and Their Management

### 7.1 Root Resorption

#### Prevalence and Risk Factors

External root resorption of teeth adjacent to impacted canines represents a significant complication, occurring in 23-48% of cases [36]. Lateral incisors are most commonly affected (60-80% of resorption cases), followed by central incisors (15-30%) and premolars (5-10%). Meta-analysis of 6 observational studies demonstrated greater root resorption in affected teeth compared to controls, with a pooled mean difference of 0.69 mm (95% CI: 0.58-0.80 mm; I-squared = 68%, tau-squared = 0.01). The prediction interval (0.47-0.91 mm) suggests consistent effect across settings.

Risk factors include:

- Mesial position of the impacted canine (sectors 4-5)
- Proximity of canine crown to adjacent root surfaces
- Prolonged traction duration
- Aggressive force application

#### Clinical Significance

Most resorption is mild (<2mm) and clinically insignificant. Severe resorption (>4mm or involving pulp) occurs in approximately 5-10% of affected teeth and may necessitate endodontic treatment or extraction [37]. The risk of severe resorption justifies baseline radiographic assessment and periodic monitoring during traction.

### 7.2 Periodontal Sequelae

#### Alveolar Bone Loss

Following successful traction, previously impacted canines exhibit reduced bone support compared to normally erupted contralateral teeth. Meta-analysis of 6 observational studies demonstrated mean bone loss of 0.51 mm (95% CI: 0.31-0.72 mm; I-squared = 85%, tau-squared = 0.03) [38]. The prediction interval (0.22-0.80 mm) indicates variability across studies. This bone loss, while statistically significant, is generally clinically acceptable with good oral hygiene.

#### Gingival Recession

Gingival recession affects approximately 10-15% of previously impacted canines, with mean recession of 0.4 mm (95% CI: 0.2-0.6 mm). Risk factors include:

- Labial impaction (versus palatal)
- Thin gingival biotype
- Inadequate keratinized tissue width
- Traction through cortical bone versus trabecular bone

#### Keratinized Tissue Width

Previously impacted canines demonstrate reduced keratinized tissue width compared to controls (mean difference −0.31 mm, 95% CI: −0.61 to −0.01 mm) [39]. When keratinized tissue is inadequate (<2mm), mucogingival surgery may be indicated.

### 7.3 Ankylosis

#### Prevalence and Diagnosis

Ankylosis—fusion of tooth root to alveolar bone—represents the most common cause of traction failure, occurring in 3.5-14.5% of surgically exposed canines [40]. Higher rates are observed in older patients and those treated with closed exposure techniques [41]. Diagnosis is suggested by:

- Failure of the tooth to move despite adequate force application (typically >3 months)
- High-pitched percussion note
- Absence of physiologic mobility
- Radiographic evidence of PDL space obliteration (though often unreliable)

CBCT provides superior detection of ankylosis through three-dimensional visualization of the bone-tooth interface [42].

#### Management

When ankylosis is confirmed, options include luxation, apicotomy, extraction, or autotransplantation [43–45]. It is critical to distinguish ankylosis from primary failure of eruption (PFE). As demonstrated in our companion review [59], PFE is characterized by a progressive posterior open bite, family history, and confirmed *PTH1R* mutations in 52-90% of cases. PFE does not respond to orthodontic force and attempting traction results in failure and iatrogenic ankylosis of adjacent teeth.

## 8. Emerging Innovations

### 8.1 Skeletal Anchorage

Temporary anchorage devices (TADs) have transformed canine traction by providing absolute anchorage independent of patient compliance [46]. Mini-implants (1.2-2.0mm diameter, 6-10mm length) placed in the premolar-molar interradicular space, palate, or infrazygomatic crest serve as stable anchor points for direct or indirect traction. Reported success rates of 88.4% for mini-screw anchorage in impacted canine extrusion exceed those of traditional dental anchorage [24]. Evidence level: Moderate–High; Clinical readiness: Established.

### 8.2 Digital Workflows and 3D Printing

Digital technologies are increasingly integrated into canine traction protocols [47]. CBCT-based planning enables precise assessment of canine position and force vector determination [48]. 3D-printed surgical guides facilitate accurate flap design and bone removal [49]. Custom CAD/CAM traction devices enable controlled force application in multiple planes [50]. Intraoral scanning can replace post-treatment CBCT once the crown is clinically visible, reducing patient radiation exposure [3]. Evidence level: Moderate; Clinical readiness: Emerging.

### 8.3 Clear Aligner-Based Traction

Clear aligner technology has been applied to canine traction through combination protocols incorporating mini-implants, elastics, and sectional wires [51]. While evidence remains limited to case series, preliminary results suggest feasibility with successful canine alignment achieved in all reported cases [3]. Evidence level: Low; Clinical readiness: Experimental.

### 8.4 Acceleration Modalities

#### Low-Level Laser Therapy (LLLT)

LLLT has been associated with shorter alignment duration in some studies, with mean differences of 58.4 days (95% CI: 28.2-88.6 days) and increased canine retraction rates of 0.27-0.31 mm/month in selected trials [52]. However, significant heterogeneity in protocols limits definitive conclusions. Current evidence suggests that while LLLT may increase the rate of tooth movement, it does not necessarily reduce total treatment duration [53]. Evidence level: Low–Moderate; Clinical readiness: Adjunct only.

#### Vibration Devices

Meta-analyses show no significant effect on alignment (mean difference 0.05 mm, 95% CI: −0.38 to 0.49 mm) and only modest effects on canine retraction (+0.27 mm/month, 95% CI: 0.19-0.35 mm) that may not be clinically meaningful [54]. Evidence level: High (negative); Clinical readiness: Not recommended.

#### Corticotomy and Micro-Osteoperforation

Surgical acceleration techniques have been associated with increased rates of tooth movement in short-term studies; however, long-term periodontal stability and cost-effectiveness require further evaluation [55]. Evidence level: Low; Clinical readiness: Limited use.

### 8.5 Genetic Testing and Personalized Medicine

The discovery that *PTH1R* mutations cause primary failure of eruption has significant implications for personalized orthodontic treatment planning [56,59]. As detailed in our companion systematic review [59], genetic testing for *PTH1R* mutations demonstrates high diagnostic utility, with a prevalence of 52-90% in PFE cases across 16 studies (487 patients). Recent meta-analytic evidence further supports the integration of genetic diagnostics into eruption disorder management algorithms [59]. Genetic testing may enable:

- Differentiation of true PFE from mechanical impaction before treatment initiation
- Identification of patients at risk for ankylosis or poor response to orthodontic force
- Informed consent regarding realistic treatment outcomes
- Selection of appropriate management strategies (traction versus prosthodontic rehabilitation)

As genetic testing becomes more accessible and affordable, integration into routine diagnostic protocols for suspected eruption disorders may become standard of care. Evidence level: Emerging; Clinical readiness: Under investigation.

Innovation evidence levels and clinical readiness are summarized in Table 9.

**Table 9:** Innovation Evidence Levels and Clinical Readiness.

| Innovation | Evidence Level | Clinical Readiness | Key Finding |
| --- | --- | --- | --- |
| **Temporary Anchorage Devices (TADs)** | ●●●○ MODERATE–<br>HIGH | **Established** | Success rates 88.4%; absolute anchorage independent of patient compliance; ideal for bilateral impaction, high anchorage demand, and non-compliant patients |
| **Digital Workflows (CBCT/3D Printing)** | ●●●○ MODERATE | **Emerging** | Improved precision in surgical planning; 3D-printed guides reduce operative time; intraoral scanning can replace post-treatment CBCT, reducing radiation exposure |
| **Clear Aligner-Based Traction** | ●○○○ LOW | **Experimental** | Feasible in selected cases when combined with mini-implants, elastics, and sectional wires; |
|  |  |  | evidence limited to case series; requires further high-quality studies |
| **Low-Level Laser Therapy (LLLT)** | ●●○○ LOW–MODERATE | **Adjunct Only** | May increase rate of tooth movement (0.27-0.31 mm/month) but does not reduce total treatment time; significant heterogeneity in protocols limits definitive conclusions |
| **Vibration Devices** | ●●●● HIGH (negative) | **Not Recommended** | No clinically significant acceleration of tooth movement (MD 0.05 mm, 95% CI: -0.38 to 0.49 mm); consistent high-quality negative evidence from meta-analyses |
| **Corticotomy/Micro-osteoperforation** | ●○○○ LOW | **Limited Use** | Short-term acceleration reported (60-100% increased rate); long-term periodontal stability and cost-effectiveness unknown; requires careful patient selection |

## 9. Evidence-Based Clinical Recommendations

### 9.1 Diagnosis and Treatment Planning

#### Recommendation 1

Comprehensive clinical and radiographic evaluation should precede any intervention. Initial assessment should include panoramic radiography with measurement of alpha-angle, d-distance, and sector classification (Grade B).

#### Recommendation 2

CBCT should be obtained when two-dimensional imaging suggests root resorption of adjacent teeth, canine position cannot be adequately visualized, surgical access planning requires three-dimensional information, or previous traction has failed (Grade B).

#### Recommendation 3

Genetic testing for *PTH1R* mutations should be considered in cases of suspected primary failure of eruption, particularly with multiple affected teeth, family history, bilateral symmetrical involvement, progressive posterior open bite, or failure to respond to orthodontic force. The diagnostic accuracy of clinical and genetic criteria for PFE is systematically evaluated in our companion review [59] (Grade C) [56,59].

### 9.2 Surgical Exposure

#### Recommendation 4

Both open and closed surgical exposure techniques are acceptable with comparable success rates. Technique selection should be based on clinician expertise, canine position, and patient factors, as outlined in the clinical decision algorithm (Figure 1) (Grade A).

#### Recommendation 5

When selecting open exposure, consider apically positioned flap techniques in areas of inadequate attached gingiva (Grade B).

#### Recommendation 6

Gold chains or stainless steel ligature wires are preferred over direct elastic attachment for bonding at surgery (Grade B).

### 9.3 Orthodontic Traction

#### Recommendation 7

Initial orthodontic forces should be light (50-75g) and directed to move the canine away from adjacent tooth roots before introducing horizontal components. Reactivate at 4-6 week intervals (Grade B).

#### Recommendation 8

Mini-implant anchorage should be considered for bilateral impaction, high anchorage demands, maximum posterior anchorage preservation, or non-compliant patients (Grade B).

#### Recommendation 9

Clear aligner-based traction protocols, when combined with mini-implants and sectional wires, represent a viable alternative in selected cases (Grade C).

### 9.4 Monitoring and Complication Management

#### Recommendation 10

Radiographic monitoring for root resorption should be performed at 6-month intervals. If resorption >2mm is detected, force modification or temporary pause should be considered (Grade B).

#### Recommendation 11

If no tooth movement is observed after 3 months of adequate force application, investigate for ankylosis and PFE through clinical assessment, percussion, CBCT, and genetic testing as per the algorithm (Figure 1) and our companion review [59] (Grade B).

#### Recommendation 12

Long-term periodontal follow-up (minimum 5 years) is recommended for previously impacted canines (Grade B).

### 9.5 Acceleration Adjuncts

#### Recommendation 13

Low-level laser therapy may be considered as a potential adjunct, though patients should be informed that the evidence base is limited and that it may not reduce total treatment time (Grade C).

#### Recommendation 14

Vibration devices are not recommended as primary acceleration modalities based on consistent high-quality negative evidence (Grade A).

#### Recommendation 15

Corticotomy and micro-osteoperforation may be considered with appropriate discussion of potential benefits versus surgical morbidity, as long-term evidence remains limited (Grade C).

### 9.6 Limitations of the Review

This review has several limitations. Included studies exhibit considerable variability in protocols, force magnitudes, and outcome definitions. Many studies fail to report exact force magnitudes or provide inadequate detail regarding force decay. For several domains—particularly acceleration modalities and clear aligner protocols—the evidence base consists primarily of case series with few randomized controlled trials. Many studies comparing open and closed exposure are retrospective and subject to selection bias. Findings are derived primarily from studies in specialized academic settings and may not fully reflect outcomes in general practice. Given the predominance of observational studies, pooled estimates should be interpreted as associations rather than causal effects.

## 10. Conclusions

Orthodontic traction of impacted canines represents a biologically complex yet clinically controllable intervention when guided by established biomechanical and diagnostic principles. This comprehensive systematic review and meta-analysis—the most extensive quantitative synthesis to date—integrates current evidence across surgical, biomechanical, radiographic, genetic, and technological domains, yielding several key conclusions.

### First, accurate diagnosis is essential prior to initiating traction

As demonstrated in our companion systematic review [59], primary failure of eruption (PFE) associated with *PTH1R* mutations must be distinguished from mechanical impaction. Application of orthodontic force to teeth affected by PFE results in failure rates of 88–98% and may induce iatrogenic ankylosis. Genetic and clinical differentiation is therefore a prerequisite to successful management.

### Second, the biomechanical foundations of canine traction are well established

Optimal force magnitudes (50–150 g), vector control relative to the center of resistance, and appropriate anchorage planning provide a rational and reproducible framework for treatment execution.

### Third, both open and closed surgical exposure techniques demonstrate high and comparable success rates

Meta-analytic synthesis indicates that technique selection may appropriately be guided by clinician expertise, spatial position of the canine, and patient-specific factors. Open exposure is associated with reduced initial traction duration and lower ankylosis risk, whereas closed exposure may offer advantages in postoperative comfort and soft-tissue aesthetics.

### Fourth, radiographic parameters enable meaningful prediction of traction duration

Alpha-angle, d-distance, and sector classification provide clinically applicable prognostic estimates. Three-dimensional analyses indicate that cusp tip displacement accounts for more than 50% of the variance in traction duration, supporting individualized treatment time forecasting.

### Fifth, complications—although relatively common—are generally mild and clinically manageable

Root resorption, alveolar bone loss, and gingival recession are typically limited in magnitude when appropriate monitoring protocols are implemented. Ankylosis, while infrequent, requires early recognition and alternative management strategies.

### Sixth, emerging innovations show variable levels of evidentiary support

Skeletal anchorage systems and digital planning workflows are supported by moderate-to-high quality evidence. Clear aligner–based traction protocols demonstrate preliminary feasibility but require controlled trials. Acceleration modalities lack sufficient high-quality evidence for routine clinical implementation.

### Collectively, the findings of this review, integrated with our companion analysis of the genetic and diagnostic foundations of PFE [**59**], support a paradigm shift toward a genetically informed, mechanistically grounded approach to all forms of failed tooth eruption

The proposed clinical decision algorithm (Figure 1) operationalizes these findings into an evidence-based management pathway.

In conclusion, orthodontic traction of impacted canines represents an established and evidence-supported clinical discipline. For the practicing orthodontist, mastery of diagnostic stratification, biomechanical control, and complication surveillance remains essential for achieving predictable functional and aesthetic outcomes.

## Supporting information

PRISMA 2020 checklist

Complete search strategies for all databases

Blank data extraction form template

Risk of bias assessments for all included studies

GRADE evidence profiles

Forest plots for all meta-analyses

Funnel plots for publication bias assessment

## Acknowledgments

The authors gratefully acknowledge the contributions of the many investigators whose work forms the foundation of this review. We thank the Ministry of Health, State of Palestine, for supporting this academic work.

## Data Availability

All data generated during this review are publicly available on the Open Science Framework project page (DOI: 10.17605/OSF.IO/3UDH6), including complete search strategies, data extraction forms, risk of bias assessments, GRADE evidence profiles, forest plots, and statistical analysis code. Study-level effect size data are provided in the supplementary materials, enabling independent verification of all meta-analyses.

**OSF Project Page:** https://osf.io/3udh6/

**OSF DOI:** https://doi.org/10.17605/OSF.IO/3UDH6

