## Supplementary material for "Canine Traction in Orthodontics: A Comprehensive Systematic Review and Meta-Analysis of Biomechanical Principles, Clinical Outcomes, and Emerging Innovations": PRISMA 2020 checklist

### **Supplementary File 1: PRISMA 2020 Checklist**

| Section and Topic | Item # | Checklist Item | Location in Manuscript |
| --- | --- | --- | --- |
| **TITLE** |  |  |  |
| Title | 1 | Identify the report as a systematic review. | Title page |
| **ABSTRACT** |  |  |  |
| Abstract | 2 | See the PRISMA 2020 for Abstracts checklist. | Abstract |
| **INTRODUCTION** |  |  |  |
| Rationale | 3 | Describe the rationale for the review in the context of existing knowledge. | Section 1.3 |
| Objectives | 4 | Provide an explicit statement of the objective(s) or question(s) the review addresses. | Section 1.3 |
| **METHODS** |  |  |  |
| Eligibility criteria | 5 | Specify the inclusion and exclusion criteria for the review. | Supplementary File 2 |
| Information sources | 6 | Specify all databases, registers, websites, organisations, reference lists and other sources searched or consulted. | Section 1.3 (Methods paragraph); Supplementary File 2 |
| Search strategy | 7 | Present the full search strategies for all databases, registers and websites. | Supplementary File 2 |
| Selection process | 8 | Specify the methods used to decide whether a study met the inclusion criteria of the review. | Supplementary File 2 |
| Data collection process | 9 | Specify the methods used to collect data from reports. | Supplementary File 3 |
| Data items | 10a | List and define all outcomes for which data were sought. | Supplementary File 3 |
|  | 10b | List and define all other variables for which data were sought. | Supplementary File 3 |
| Study risk of bias assessment | 11 | Specify the methods used to assess risk of bias in the included studies. | Supplementary File 4 |
| Effect measures | 12 | Specify for each outcome the effect measure(s) used in the synthesis. | Throughout manuscript |
| Synthesis methods | 13a | Describe the processes used to decide which studies were eligible for each synthesis. | Section 1.3 |
|  | 13b | Describe any methods required to prepare the data for presentation or synthesis. | Section 1.3 |
|  | 13c | Describe any methods used to tabulate or visually display results of individual studies and syntheses. | Supplementary Files 6, 7 |
|  | 13d | Describe any methods used to synthesize results and provide a rationale for the choice(s). | Section 1.3 |
|  | 13e | Describe any methods used to explore possible causes of heterogeneity among study results. | Supplementary File 6 |
|  | 13f | Describe any sensitivity analyses conducted to assess robustness of the synthesized results. | Supplementary File 6 |
| Reporting bias assessment | 14 | Describe any methods used to assess risk of bias due to missing results in a synthesis. | Supplementary File 7 |
| Certainty assessment | 15 | Describe any methods used to assess certainty (or confidence) in the body of evidence for an outcome. | Supplementary File 5 |
| **RESULTS** |  |  |  |
| Study selection | 16a | Describe the results of the search and selection process, from the number of records identified in the search to the number of studies included in the review. | Section 1.3 (Methods paragraph) |
|  | 16b | Cite studies that might appear to meet the inclusion criteria, but which were excluded, and explain why they were excluded. | N/A |
| Study characteristics | 17 | Cite each included study and present its characteristics. | Throughout manuscript (citations) |
| Risk of bias in studies | 18 | Present assessments of risk of bias for each included study. | Supplementary File 4 |
| Results of individual studies | 19 | For all outcomes, present, for each study, summary statistics for each group and an effect estimate. | Throughout manuscript |
| Results of syntheses | 20a | For each synthesis, briefly summarise the characteristics and risk of bias among contributing studies. | Supplementary Files 4, 5 |
|  | 20b | Present results of all statistical syntheses conducted. | Throughout manuscript |
|  | 20c | Present results of any investigations of possible causes of heterogeneity. | Supplementary File 6 |
|  | 20d | Present results of any sensitivity analyses conducted to assess robustness of the synthesized results. | Supplementary File 6 |
| Reporting biases | 21 | Present assessments of risk of bias due to missing results for each synthesis assessed. | Supplementary File 7 |
| Certainty of evidence | 22 | Present assessments of certainty (or confidence) in the body of evidence for each outcome assessed. | Supplementary File 5 |
| **DISCUSSION** |  |  |  |
| Discussion | 23a | Provide a general interpretation of the results in the context of other evidence. | Section 9 |
|  | 23b | Discuss any limitations of the evidence included in the review. | Section 8.6 |
|  | 23c | Discuss any limitations of the review processes used. | Section 8.6 |
|  | 23d | Discuss implications of the results for practice, policy, and future research. | Sections 8, 9 |
| **OTHER INFORMATION** |  |  |  |
| Registration and protocol | 24a | Provide registration information for the review, including register name and registration number. | Title page |
|  | 24b | Indicate where the review protocol can be accessed. | Title page |
|  | 24c | Describe and explain any amendments to information provided at registration or in the protocol. | N/A |
| Support | 25 | Describe sources of financial or non-financial support for the review, and the role of the funders or sponsors in the review. | Funding statement |
| Competing interests | 26 | Declare any competing interests of review authors. | Conflicts of Interest |
| Availability of data, code and other materials | 27 | Report which of the following are publicly available and where they can be found: template data collection forms; data extracted from included studies; data used for all analyses; analytic code; any other materials used in the review. | Supplementary Files 1-7; Open Science Framework |
