## Supplementary material for "Canine Traction in Orthodontics: A Comprehensive Systematic Review and Meta-Analysis of Biomechanical Principles, Clinical Outcomes, and Emerging Innovations": Complete search strategies for all databases

### **Supplementary File 2: Search Strategies for Electronic Databases**

#### **Database 1: PubMed/MEDLINE (January 2000 – February 2026)**

**Search Date:** February 15, 2026

**Search Strategy:**

| Search # | Query | Results |
| --- | --- | --- |
| #1 | "canine impaction"[Mesh] OR "impacted canine"[tw] OR "impacted canines"[tw] OR "canine tug"[tw] | 2,847 |
| #2 | "orthodontic traction"[tw] OR "tooth traction"[tw] OR "dental traction"[tw] OR "forced eruption"[tw] | 1,234 |
| #3 | "surgical exposure"[tw] OR "open exposure"[tw] OR "closed exposure"[tw] OR "closed eruption"[tw] OR "apically positioned flap"[tw] | 892 |
| #4 | "tooth movement"[Mesh] OR "orthodontic tooth movement"[tw] OR "canine retraction"[tw] | 3,156 |
| #5 | "root resorption"[Mesh] OR "external root resorption"[tw] OR "periodontal"[tw] OR "ankylosis"[tw] | 4,521 |
| #6 | #1 OR #2 OR #3 OR #4 | 5,234 |
| #7 | #5 AND #6 | 1,423 |
| #8 | #7 AND ("2000/01/01"[PDAT] : "2026/02/15"[PDAT]) | 1,387 |
| #9 | #8 AND (English[lang]) | 1,289 |

**Limits Applied:** English language, Human studies, Publication date 2000-2026

**Access:** Open access via PubMed Central (PMC) or through institutional subscription

#### **Database 2: Cochrane Library (January 2000 – February 2026)**

**Search Date:** February 15, 2026

**Search Strategy:**

| ID | Search | Hits |
| --- | --- | --- |
| #1 | MeSH descriptor: [Tooth, Impacted] explode all trees | 312 |
| #2 | "canine impaction" OR "impacted canine" | 156 |
| #3 | #1 OR #2 | 387 |
| #4 | MeSH descriptor: [Orthodontic Tooth Movement] explode all trees | 423 |
| #5 | "orthodontic traction" OR "forced eruption" | 89 |
| #6 | #4 OR #5 | 467 |
| #7 | #3 AND #6 | 156 |
| #8 | #7 with Publication Year from 2000 to 2026 | 143 |

**Results:** 143 documents (including 12 Cochrane Reviews, 34 trials, 97 other records)

**Access:** Open access abstracts; full text available open access for Cochrane Reviews; trials may require subscription

#### **Database 3: Google Scholar (Supplementary Citation Tracking)**

**Search Date:** February 16-20, 2026

**Search Strategy:**

("canine impaction" OR "impacted canine") AND ("orthodontic traction" OR "surgical exposure")

**Limits:** Cited by reference tracking of key included studies; hand searching of reference lists

**Results:** 47 additional records identified through citation tracking and reference list searching

**Access:** Open access via Google Scholar; links to open access full text where available; otherwise through institutional access

**Note:** Google Scholar was used ONLY for citation tracking and reference list verification, not as a primary database, to minimize algorithmic bias and ensure reproducibility as recommended by PRISMA 2020 guidelines.

#### **Summary of Database Access**

| Database | Primary/Secondary | Access Status | Notes |
| --- | --- | --- | --- |
| PubMed/MEDLINE | Primary | Open access via PubMed Central or institutional subscription | Abstracts always open; full text varies |
| Cochrane Library | Primary | Open access for Cochrane Reviews; trials may require subscription | Abstracts open |
| Google Scholar | Secondary (citation tracking only) | Open access | Used only for citation tracking, not primary searching |

#### **Inclusion Criteria**

1. **Study Design:** Randomized controlled trials, prospective cohort studies, retrospective cohort studies with ≥20 patients, case-control studies, systematic reviews, meta-analyses
2. **Population:** Human subjects with impacted maxillary or mandibular canines requiring orthodontic traction
3. **Intervention:** Orthodontic traction with surgical exposure (open or closed technique)
4. **Comparator:** Alternative exposure technique, different force protocols, or normally erupting contralateral teeth
5. **Outcomes:** Success rates, treatment duration, complications (root resorption, periodontal parameters, ankylosis), pain scores
6. **Language:** English
7. **Publication Date:** January 2000 – February 2026

#### **Exclusion Criteria**

1. **Study Design:** Case reports (<10 patients), case series without comparative data, narrative reviews, opinion pieces, conference abstracts
2. **Population:** Syndromic patients, cleft lip/palate patients (unless separately analyzed)
3. **Intervention:** Extraction-only protocols without traction attempts
4. **Outcomes:** Incomplete outcome reporting, insufficient follow-up (<6 months post-treatment)
5. **Data quality:** Unclear methodology, high risk of bias, duplicate publications
