## Supplementary material for "Canine Traction in Orthodontics: A Comprehensive Systematic Review and Meta-Analysis of Biomechanical Principles, Clinical Outcomes, and Emerging Innovations": Blank data extraction form template

### **Supplementary File 3: Data Extraction Form**

#### **Study Information**

| Field | Data Entry |
| --- | --- |
| Study ID | [First Author Year] |
| Title |  |
| Journal |  |
| Publication Year |  |
| Country |  |
| Study Design | □ RCT □ Prospective cohort □ Retrospective cohort □ Case-control □ Systematic review □ Meta-analysis |
| Setting | □ University □ Hospital □ Private practice □ Multi-center |
| Funding Source |  |
| Conflict of Interest |  |

#### **Patient Characteristics**

| Field | Data Entry |
| --- | --- |
| Sample Size (n) |  |
| Age Range (years) |  |
| Mean Age (SD) |  |
| Sex (M/F) |  |
| Unilateral/Bilateral |  |
| Impaction Location | □ Palatal □ Buccal □ Mid-alveolar □ Mandibular |
| Side Distribution | □ Right □ Left |
| Follow-up Duration |  |

#### **Intervention Details (Surgical Exposure)**

| Field | Open Exposure | Closed Exposure |
| --- | --- | --- |
| Technique Description |  |  |
| Flap Design |  |  |
| Bone Removal Method |  |  |
| Attachment Type |  |  |
| Bonding Material |  |  |
| Periodontal Pack | □ Yes □ No | □ Yes □ No |
| Time to Traction Start |  |  |

#### **Intervention Details (Orthodontic Traction)**

| Field | Data Entry |
| --- | --- |
| Force Type | □ Elastomeric chain □ NiTi spring □ Superelastic wire □ Other |
| Force Magnitude (g) |  |
| Force Direction | □ Vertical □ Distal □ Buccal □ Combination |
| Activation Interval |  |
| Anchorage Type | □ Dental □ TAD □ Headgear □ Combination |
| Appliance Type | □ Fixed □ Removable □ Clear aligner |

#### **Outcomes: Success Rates**

| Outcome | Open Exposure | Closed Exposure | Control | P-value |
| --- | --- | --- | --- | --- |
| Success Rate (n/N) |  |  |  |  |
| Success Rate (%) [95% CI] |  |  |  |  |
| Definition of Success |  |  |  |  |

#### **Outcomes: Treatment Duration (months)**

| Outcome | Open Exposure | Closed Exposure | Control | Mean Difference [95% CI] |
| --- | --- | --- | --- | --- |
| Time to Eruption |  |  |  |  |
| Active Traction Time |  |  |  |  |
| Total Treatment Time |  |  |  |  |

#### **Outcomes: Root Resorption**

| Outcome | Affected Teeth | Unaffected Teeth | Mean Difference [95% CI] |
| --- | --- | --- | --- |
| Prevalence (%) |  |  |  |
| Central Incisors (mm) |  |  |  |
| Lateral Incisors (mm) |  |  |  |
| Canine (mm) |  |  |  |

#### **Outcomes: Periodontal Parameters**

| Parameter | Affected Teeth | Control Teeth | Mean Difference [95% CI] |
| --- | --- | --- | --- |
| Probing Depth (mm) |  |  |  |
| Keratinized Tissue Width (mm) |  |  |  |
| Gingival Recession (mm) |  |  |  |
| Bone Loss (mm) |  |  |  |

#### **Outcomes: Ankylosis**

| Outcome | Open Exposure | Closed Exposure | Total | P-value |
| --- | --- | --- | --- | --- |
| Ankylosis Rate (n/N) |  |  |  |  |
| Ankylosis Rate (%) |  |  |  |  |
| Time to Diagnosis (months) |  |  |  |  |

#### **Outcomes: Pain and Patient-Reported Outcomes**

| Outcome | Open Exposure | Closed Exposure | Mean Difference [95% CI] |
| --- | --- | --- | --- |
| Pain Score (VAS 0-10) |  |  |  |
| Analgesic Use (%) |  |  |  |
| Functional Impairment |  |  |  |
| Patient Satisfaction |  |  |  |

#### **Risk of Bias Assessment**

| Domain | Rating | Comments |
| --- | --- | --- |
| Selection Bias | □ Low □ High □ Unclear |  |
| Performance Bias | □ Low □ High □ Unclear |  |
| Detection Bias | □ Low □ High □ Unclear |  |
| Attrition Bias | □ Low □ High □ Unclear |  |
| Reporting Bias | □ Low □ High □ Unclear |  |
| Overall | □ Low □ High □ Unclear |  |

#### **Notes**

| Field | Notes |
| --- | --- |
| Key Findings |  |
| Limitations |  |
| Correspondence |  |
