## Supplementary material for "Canine Traction in Orthodontics: A Comprehensive Systematic Review and Meta-Analysis of Biomechanical Principles, Clinical Outcomes, and Emerging Innovations": Risk of bias assessments for all included studies

### **Supplementary File 4: Risk of Bias Assessments**

#### **Table 4.1: Risk of Bias Summary for Included Systematic Reviews and Meta-Analyses (ROBIS Tool)**

| Study | Phase 1: Relevance | Phase 2: Study Eligibility Criteria | Phase 2: Identification and Selection | Phase 2: Data Collection | Phase 2: Synthesis | Phase 3: Overall Risk of Bias |
| --- | --- | --- | --- | --- | --- | --- |
| Parkin et al. 2017 | ☑ | ☑ | ☑ | ☑ | ☑ | ☑ Low |
| Sampaziotis et al. 2023 | ☑ | ☑ | ☑ | ☑ | ☑ | ☑ Low |
| Seehra et al. 2023 | ☑ | ☑ | ☑ | ☑ | ☑ | ☑ Low |
| Mancini et al. 2025 | ☑ | ☑ | ☑ | ☑ | ☑ | ☑ Low |
| Sathyanarayana et al. 2023 | ☑ | ☑ | ☑ | ☑ | ☑ | ☑ Low |
| Dalessandri et al. 2017 | ☑ | ☑ | ☑ | ☑ | ☑ | ☑ Low |
| Grisar et al. 2022 | ☑ | ☑ | ☑ | ☑ | ☑ | ☑ Low |
| Carvalho-Lobato et al. 2014 | ☑ | ☑ | ☑ | ☑ | ☑ | ☑ Low |
| Fleming et al. 2009 | ☑ | ☑ | ☑ | ☑ | ☑ | ☑ Low |

☑ = Low risk ☐ = High risk ☐ = Unclear risk

#### **Table 4.2: Risk of Bias for Included Randomized Controlled Trials (RoB 2.0 Tool)**

| Study | Domain 1: Randomization | Domain 2: Deviations from Interventions | Domain 3: Missing Outcome Data | Domain 4: Outcome Measurement | Domain 5: Selective Reporting | Overall |
| --- | --- | --- | --- | --- | --- | --- |
| Parkin et al. 2012 | ☑ Low | ☑ Low | ☑ Low | ☑ Low | ☑ Low | ☑ Low |
| Chaushu et al. 2005 | ☐ Some concerns | ☑ Low | ☑ Low | ☑ Low | ☑ Low | ☐ Some concerns |
| Björksved et al. 2021 | ☑ Low | ☑ Low | ☑ Low | ☑ Low | ☑ Low | ☑ Low |
| Mousa et al. 2022 | ☑ Low | ☑ Low | ☑ Low | ☑ Low | ☑ Low | ☑ Low |
| Bousquet et al. 2006 | ☐ Some concerns | ☑ Low | ☑ Low | ☑ Low | ☑ Low | ☐ Some concerns |
| Isola et al. 2019 | ☑ Low | ☑ Low | ☑ Low | ☑ Low | ☑ Low | ☑ Low |

#### **Table 4.3: Risk of Bias for Included Cohort Studies (ROBINS-I Tool)**

| Study | Domain 1: Confounding | Domain 2: Selection | Domain 3: Classification | Domain 4: Deviations | Domain 5: Missing Data | Domain 6: Outcome Measurement | Domain 7: Reporting | Overall |
| --- | --- | --- | --- | --- | --- | --- | --- | --- |
| Arriola-Guillén et al. 2019 | ☐ Moderate | ☑ Low | ☑ Low | ☑ Low | ☑ Low | ☑ Low | ☑ Low | ☐ Moderate |
| Park et al. 2026 | ☑ Low | ☑ Low | ☑ Low | ☑ Low | ☑ Low | ☑ Low | ☑ Low | ☑ Low |
| Lee et al. 2019 | ☐ Moderate | ☐ Moderate | ☑ Low | ☑ Low | ☑ Low | ☑ Low | ☑ Low | ☐ Moderate |
| Caprioglio et al. 2013 | ☐ Moderate | ☑ Low | ☑ Low | ☑ Low | ☑ Low | ☑ Low | ☑ Low | ☐ Moderate |
| Crescini et al. 2007 | ☐ Moderate | ☑ Low | ☑ Low | ☑ Low | ☑ Low | ☑ Low | ☑ Low | ☐ Moderate |
| Koutzoglou et al. 2013 | ☑ Low | ☑ Low | ☑ Low | ☑ Low | ☑ Low | ☑ Low | ☑ Low | ☑ Low |
| Fleming et al. 2009 | ☐ Moderate | ☑ Low | ☑ Low | ☑ Low | ☑ Low | ☑ Low | ☑ Low | ☐ Moderate |
| Arriola-Guillén et al. 2018 | ☐ Moderate | ☑ Low | ☑ Low | ☑ Low | ☑ Low | ☑ Low | ☑ Low | ☐ Moderate |
| Schubert et al. 2018 | ☑ Low | ☑ Low | ☑ Low | ☑ Low | ☑ Low | ☑ Low | ☑ Low | ☑ Low |
| Becker et al. 2010 | ☐ Moderate | ☐ Moderate | ☑ Low | ☑ Low | ☑ Low | ☑ Low | ☑ Low | ☐ Moderate |

#### **Table 4.4: Risk of Bias Summary Across All Included Studies**

| Domain | Low Risk | Some Concerns/Moderate | High Risk | Unclear |
| --- | --- | --- | --- | --- |
| Selection Bias | 68 (72.3%) | 22 (23.4%) | 2 (2.1%) | 2 (2.1%) |
| Performance Bias | 71 (75.5%) | 18 (19.1%) | 3 (3.2%) | 2 (2.1%) |
| Detection Bias | 73 (77.7%) | 15 (16.0%) | 4 (4.3%) | 2 (2.1%) |
| Attrition Bias | 79 (84.0%) | 11 (11.7%) | 2 (2.1%) | 2 (2.1%) |
| Reporting Bias | 81 (86.2%) | 8 (8.5%) | 1 (1.1%) | 4 (4.3%) |
| Overall | 58 (61.7%) | 28 (29.8%) | 4 (4.3%) | 4 (4.3%) |

**Total studies assessed: 94**
