## Supplementary material for "Canine Traction in Orthodontics: A Comprehensive Systematic Review and Meta-Analysis of Biomechanical Principles, Clinical Outcomes, and Emerging Innovations": GRADE evidence profiles

### **Supplementary File 5: GRADE Evidence Profiles**

#### **Table 5.1: GRADE Profile – Open vs. Closed Surgical Exposure for Impacted Canines**

| Outcome | № of Studies | Study Design | Risk of Bias | Inconsistency | Indirectness | Imprecision | Other Considerations | Certainty |
| --- | --- | --- | --- | --- | --- | --- | --- | --- |
| **Success Rate** | 12 | RCTs & observational | Not serious¹ | Not serious | Not serious | Not serious | Strong association | ⚫⚫⚪⚪ MODERATE |
| **Treatment Duration** | 9 | RCTs & observational | Serious² | Serious³ | Not serious | Not serious | None | ⚫⚫⚪⚪ MODERATE |
| **Ankylosis Risk** | 6 | Observational | Serious² | Not serious | Not serious | Serious⁴ | Dose-response gradient | ⚫⚪⚪⚪ LOW |
| **Postoperative Pain** | 5 | RCTs & observational | Not serious¹ | Not serious | Not serious | Not serious | Large effect | ⚫⚫⚫⚪ HIGH |
| **Root Resorption** | 8 | Observational | Serious² | Not serious | Not serious | Not serious | None | ⚫⚫⚪⚪ MODERATE |
| **Periodontal Outcomes** | 11 | Observational | Serious² | Not serious | Not serious | Not serious | None | ⚫⚫⚪⚪ MODERATE |

**Explanations:**
¹ Some studies with moderate risk of bias, but sensitivity analysis excluding them did not change effect estimates
² Primarily observational studies with inherent confounding
³ Wide prediction intervals indicating substantial variability
⁴ Confidence interval includes clinically unimportant effects

#### **Table 5.2: GRADE Profile – Radiographic Predictors of Traction Duration**

| Outcome | № of Studies | Study Design | Risk of Bias | Inconsistency | Indirectness | Imprecision | Other Considerations | Certainty |
| --- | --- | --- | --- | --- | --- | --- | --- | --- |
| **α-Angle** | 14 | Observational | Serious¹ | Not serious | Not serious | Not serious | Strong association | ⚫⚫⚪⚪ MODERATE |
| **d-Distance** | 12 | Observational | Serious¹ | Not serious | Not serious | Not serious | Strong association, dose-response | ⚫⚫⚫⚪ HIGH |
| **Sector Location** | 11 | Observational | Serious¹ | Not serious | Not serious | Not serious | Strong association | ⚫⚫⚪⚪ MODERATE |
| **3D Cusp Displacement** | 3 | Observational | Not serious² | Not serious | Not serious | Serious³ | None | ⚫⚫⚪⚪ MODERATE |

**Explanations:**
¹ Primarily retrospective cohort studies
² Recent prospective studies with CBCT validation
³ Limited number of studies, though effect size consistent

#### **Table 5.3: GRADE Profile – Complications of Canine Traction**

| Outcome | № of Studies | Study Design | Risk of Bias | Inconsistency | Indirectness | Imprecision | Other Considerations | Certainty |
| --- | --- | --- | --- | --- | --- | --- | --- | --- |
| **Root Resorption (adjacent incisors)** | 16 | Observational | Serious¹ | Not serious | Not serious | Not serious | Strong association | ⚫⚫⚪⚪ MODERATE |
| **Alveolar Bone Loss** | 9 | Observational | Serious¹ | Not serious | Not serious | Not serious | None | ⚫⚫⚪⚪ MODERATE |
| **Gingival Recession** | 8 | Observational | Serious¹ | Not serious | Not serious | Not serious | None | ⚫⚫⚪⚪ MODERATE |
| **Ankylosis** | 7 | Observational | Serious¹ | Not serious | Not serious | Serious² | None | ⚫⚪⚪⚪ LOW |

**Explanations:**
¹ Primarily observational studies with potential selection bias
² Confidence intervals wide due to relatively rare outcome

#### **Table 5.4: GRADE Profile – Acceleration Modalities**

| Outcome | № of Studies | Study Design | Risk of Bias | Inconsistency | Indirectness | Imprecision | Other Considerations | Certainty |
| --- | --- | --- | --- | --- | --- | --- | --- | --- |
| **Low-Level Laser Therapy** | 5 | RCTs & observational | Serious¹ | Serious² | Not serious | Serious³ | None | ⚫⚪⚪⚪ VERY LOW |
| **Vibration Devices** | 4 | RCTs | Not serious | Serious² | Not serious | Serious³ | No effect | ⚫⚫⚪⚪ LOW |
| **Corticotomy/Micro-osteoperforation** | 6 | Observational | Serious¹ | Serious² | Not serious | Serious³ | None | ⚫⚪⚪⚪ VERY LOW |

**Explanations:**
¹ Limited RCT evidence, primarily case series
² Significant heterogeneity in protocols and outcome measures
³ Small sample sizes, wide confidence intervals

#### **Table 5.5: GRADE Profile – Clear Aligner-Based Traction**

| Outcome | № of Studies | Study Design | Risk of Bias | Inconsistency | Indirectness | Imprecision | Other Considerations | Certainty |
| --- | --- | --- | --- | --- | --- | --- | --- | --- |
| **Success Rate** | 3 | Observational | Serious¹ | Not serious | Not serious | Serious² | None | ⚫⚪⚪⚪ VERY LOW |
| **Treatment Duration** | 2 | Observational | Serious¹ | Not serious | Not serious | Serious² | None | ⚫⚪⚪⚪ VERY LOW |
| **Radiation Reduction** | 2 | Observational | Serious¹ | Not serious | Not serious | Not serious³ | Large potential benefit | ⚫⚫⚪⚪ MODERATE |

**Explanations:**
¹ Limited number of studies, all case series
² Small sample sizes
³ Consistent finding across studies
