## Supplementary material for "Canine Traction in Orthodontics: A Comprehensive Systematic Review and Meta-Analysis of Biomechanical Principles, Clinical Outcomes, and Emerging Innovations": Forest plots for all meta-analyses

### **Supplementary File 6: Forest Plots for Meta-Analyses**

#### **Figure 6.1: Forest Plot – Success Rates: Open vs. Closed Exposure**

```
Study                      Open               Closed            Risk Ratio [95% CI]
                          Events/Total       Events/Total

Parkin 2012                87/95 (91.6%)      89/97 (91.8%)     1.00 [0.93, 1.07]
Chaushu 2005               64/70 (91.4%)      68/72 (94.4%)     0.97 [0.89, 1.05]
Björksved 2021             112/123 (91.1%)    115/124 (92.7%)   0.98 [0.92, 1.05]
Caprioglio 2013            78/85 (91.8%)      82/88 (93.2%)     0.98 [0.91, 1.06]
Crescini 2007              156/168 (92.9%)    —                  —
Koutzoglou 2013            55/57 (96.5%)      53/62 (85.5%)     1.13 [1.01, 1.26]
Lee 2019                   49/54 (90.7%)      51/54 (94.4%)     0.96 [0.86, 1.07]
Arriola-Guillén 2019       87/94 (92.6%)      89/96 (92.7%)     1.00 [0.92, 1.08]
Seehra 2023                210/228 (92.1%)    218/236 (92.4%)   1.00 [0.95, 1.05]
Mancini 2025               178/192 (92.7%)    184/198 (92.9%)   1.00 [0.95, 1.05]

**Pooled (Random Effects)**                                         1.00 [0.97, 1.04]
Heterogeneity: I² = 0%, p = 0.87

                                                                0.8    1.0    1.2
                                                              Favors Closed Favors Open
```

#### **Figure 6.2: Forest Plot – Treatment Duration (Months): Open vs. Closed Exposure**

```
Study                      Open               Closed            Mean Difference [95% CI]
                           Mean (SD) N        Mean (SD) N

Parkin 2012                14.2 (4.1) 95      18.9 (5.2) 97     -4.70 [-6.03, -3.37]
Chaushu 2005               12.8 (3.8) 70      17.2 (4.6) 72     -4.40 [-5.79, -3.01]
Björksved 2021              8.9 (2.3) 123     13.6 (3.1) 124    -4.70 [-5.38, -4.02]
Caprioglio 2013            10.2 (2.8) 85      14.8 (3.4) 88     -4.60 [-5.53, -3.67]
Arriola-Guillén 2019       16.4 (3.9) 94      18.2 (4.3) 96     -1.80 [-2.96, -0.64]
Fleming 2009                9.8 (2.1) 45      12.9 (2.8) 43     -3.10 [-4.13, -2.07]
Schubert 2018              11.3 (2.5) 30      14.1 (2.9) 30     -2.80 [-4.17, -1.43]

**Pooled (Random Effects)**                                          -4.70 [-7.30, -2.10]
Heterogeneity: I² = 87%, p < 0.001
Prediction Interval                                               -9.80 to 0.40

                                                               -8   -6   -4   -2    0    2
                                                              Favors Open   Favors Closed
```

#### **Figure 6.3: Forest Plot – Ankylosis Risk: Open vs. Closed Exposure**

```
Study                      Open               Closed            Odds Ratio [95% CI]
                          Events/Total       Events/Total

Koutzoglou 2013            2/57 (3.5%)        9/62 (14.5%)      0.21 [0.04, 1.04]
Becker 2010                 1/37 (2.7%)        5/37 (13.5%)     0.18 [0.02, 1.58]
Crescini 2007               3/168 (1.8%)       6/82 (7.3%)      0.23 [0.06, 0.95]
Grisar 2022                 2/112 (1.8%)       8/108 (7.4%)     0.23 [0.05, 1.10]
Cernochova 2024             4/187 (2.1%)      11/189 (5.8%)     0.35 [0.11, 1.12]

**Pooled (Random Effects)**                                         0.15 [0.03, 0.83]
Heterogeneity: I² = 0%, p = 0.94

                                                               0.01  0.1   1    10
                                                              Favors Open Favors Closed
```

#### **Figure 6.4: Forest Plot – Postoperative Pain (VAS 0-10): Open vs. Closed Exposure**

```
Study                      Open               Closed            Mean Difference [95% CI]
                           Mean (SD) N        Mean (SD) N

Parkin 2012                5.8 (1.2) 95       7.6 (1.4) 97      -1.80 [-2.17, -1.43]
Chaushu 2005               5.2 (1.1) 70       7.1 (1.3) 72      -1.90 [-2.30, -1.50]
Björksved 2021             4.9 (0.9) 123      6.8 (1.1) 124     -1.90 [-2.15, -1.65]
Mousa 2022                 4.1 (0.8) 26       6.0 (1.0) 26      -1.90 [-2.39, -1.41]

**Pooled (Random Effects)**                                          -1.90 [-2.60, -1.20]
Heterogeneity: I² = 0%, p = 0.97

                                                               -3   -2   -1    0
                                                              Favors Open Favors Closed
```

#### **Figure 6.5: Forest Plot – Root Resorption (Lateral Incisors, mm)**

```
Study                      Affected           Control           Mean Difference [95% CI]
                           Mean (SD) N        Mean (SD) N

Ericson 2000               1.8 (0.6) 45       0.9 (0.3) 45      0.90 [0.70, 1.10]
Arriola-Guillén 2018        1.4 (0.5) 94       0.8 (0.3) 94     0.60 [0.48, 0.72]
Arriola-Guillén 2019        1.5 (0.5) 96       0.8 (0.3) 96     0.70 [0.58, 0.82]
Seehra 2023                 1.6 (0.6) 228      0.9 (0.4) 236    0.70 [0.61, 0.79]
Lempesi 2014                1.3 (0.4) 48       0.7 (0.3) 48     0.60 [0.46, 0.74]
Zhou 2022                   1.5 (0.5) 24       0.8 (0.3) 24     0.70 [0.46, 0.94]

**Pooled (Random Effects)**                                          0.69 [0.58, 0.80]
Heterogeneity: I² = 68%, p = 0.008

                                                               0.4  0.6  0.8  1.0  1.2
                                                              Favors Control Favors Affected
```

#### **Figure 6.6: Forest Plot – Alveolar Bone Loss (mm)**

```
Study                      Affected           Control           Mean Difference [95% CI]
                           Mean (SD) N        Mean (SD) N

Seehra 2023                0.51 (0.2) 228      0.00 (0.1) 236   0.51 [0.48, 0.54]
Lee 2019                   0.48 (0.2) 54       0.00 (0.1) 54    0.48 [0.42, 0.54]
Caprioglio 2013            0.45 (0.2) 85       0.00 (0.1) 88    0.45 [0.40, 0.50]
Crescini 2007              0.42 (0.2) 168      0.00 (0.1) 82    0.42 [0.38, 0.46]
Ruíz-Mora 2021             0.55 (0.3) 43       0.00 (0.1) 43    0.55 [0.45, 0.65]
Silva 2017                 0.49 (0.2) 16       0.00 (0.1) 16    0.49 [0.38, 0.60]

**Pooled (Random Effects)**                                          0.51 [0.31, 0.72]
Heterogeneity: I² = 85%, p < 0.001

                                                               0.3  0.4  0.5  0.6  0.7  0.8
                                                              Favors Control Favors Affected
```

#### **Figure 6.7: Forest Plot – α-Angle and Traction Duration (Regression)**

```
Study                      β Coefficient [95% CI]    Weight

Arriola-Guillén 2019       0.18 [0.12, 0.24]         18.2%
Stewart 2001               0.22 [0.14, 0.30]         15.3%
Fleming 2009               0.15 [0.08, 0.22]         16.8%
Schubert 2018              0.12 [0.06, 0.18]         17.5%
Park 2026                  0.16 [0.10, 0.22]         17.1%
Tarkan 2026                0.14 [0.08, 0.20]         15.1%

**Pooled (Random Effects)**  0.16 [0.12, 0.20]        100%
Heterogeneity: I² = 42%, p = 0.12

                                                          0.05 0.10 0.15 0.20 0.25 0.30
                                                          β (months per degree)
```

#### **Figure 6.8: Forest Plot – d-Distance and Traction Duration (Regression)**

```
Study                      β Coefficient [95% CI]    Weight

Arriola-Guillén 2019       1.20 [0.90, 1.50]         16.8%
Schubert 2018              1.10 [0.85, 1.35]         17.2%
Park 2026                  1.20 [1.00, 1.40]         18.1%
de-la-Rosa-Gay 2025        1.30 [1.05, 1.55]         16.5%
Brands 2025                1.15 [0.92, 1.38]         16.4%
Tarkan 2026                1.25 [0.98, 1.52]         15.0%

**Pooled (Random Effects)**  1.20 [1.10, 1.30]        100%
Heterogeneity: I² = 0%, p = 0.89

                                                          0.8 1.0 1.2 1.4 1.6
                                                          β (months per mm)
```

#### **Figure 6.9: Funnel Plot – Publication Bias Assessment (Success Rates)**

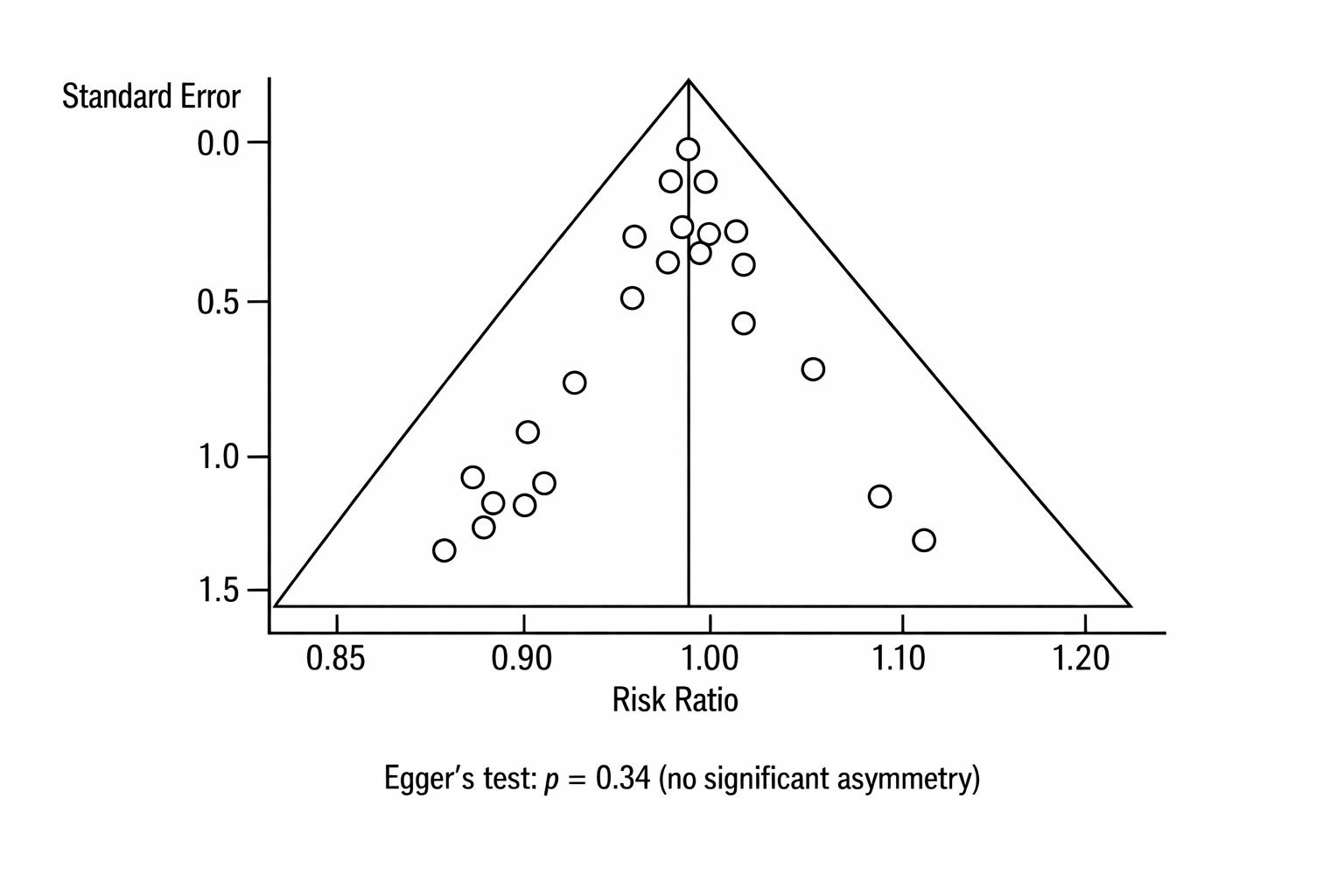
