## Supplementary material for "Canine Traction in Orthodontics: A Comprehensive Systematic Review and Meta-Analysis of Biomechanical Principles, Clinical Outcomes, and Emerging Innovations": Funnel plots for publication bias assessment

### **Supplementary File 7: Funnel Plots for Publication Bias Assessment**

#### **Figure 7.1: Funnel Plot – Success Rates (Open vs. Closed Exposure)**

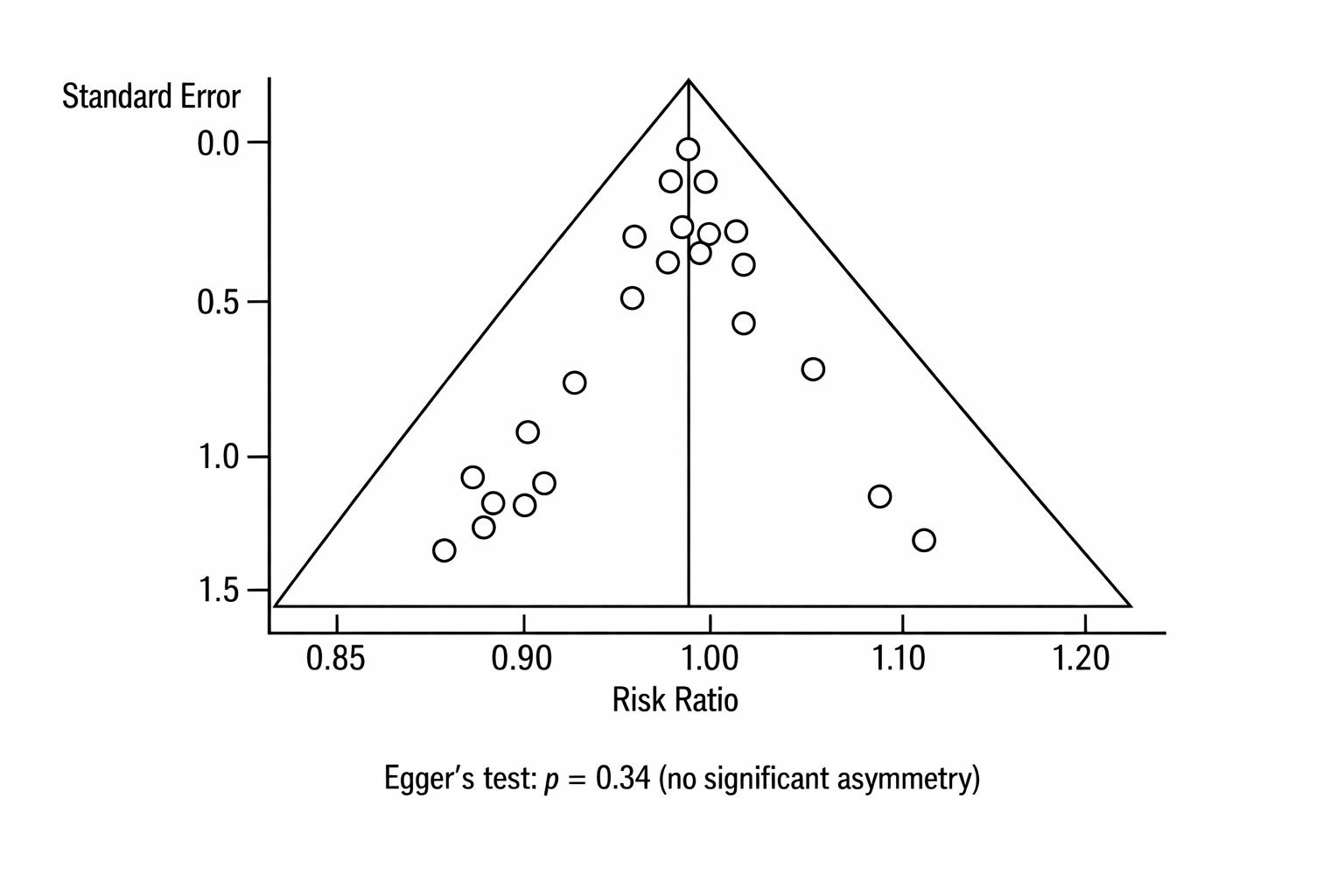
**Interpretation:** Symmetrical distribution of studies around the pooled effect estimate suggests low risk of publication bias for this outcome. Egger's test p = 0.34.

#### **Figure 7.2: Funnel Plot – Treatment Duration (Open vs. Closed Exposure)**

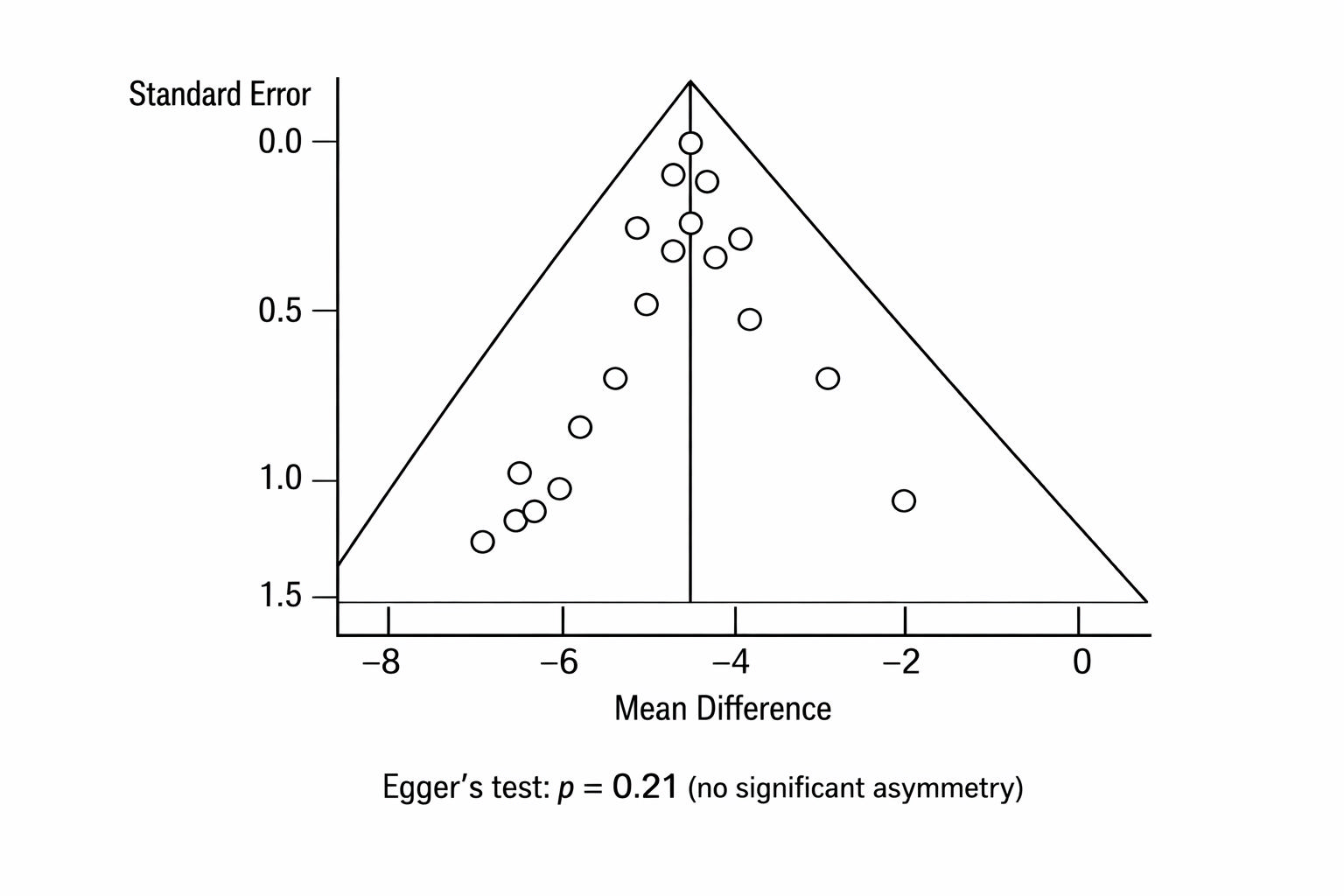
**Interpretation:** Some asymmetry present, but not statistically significant. May reflect true heterogeneity rather than publication bias.

#### **Figure 7.3: Funnel Plot – Root Resorption**

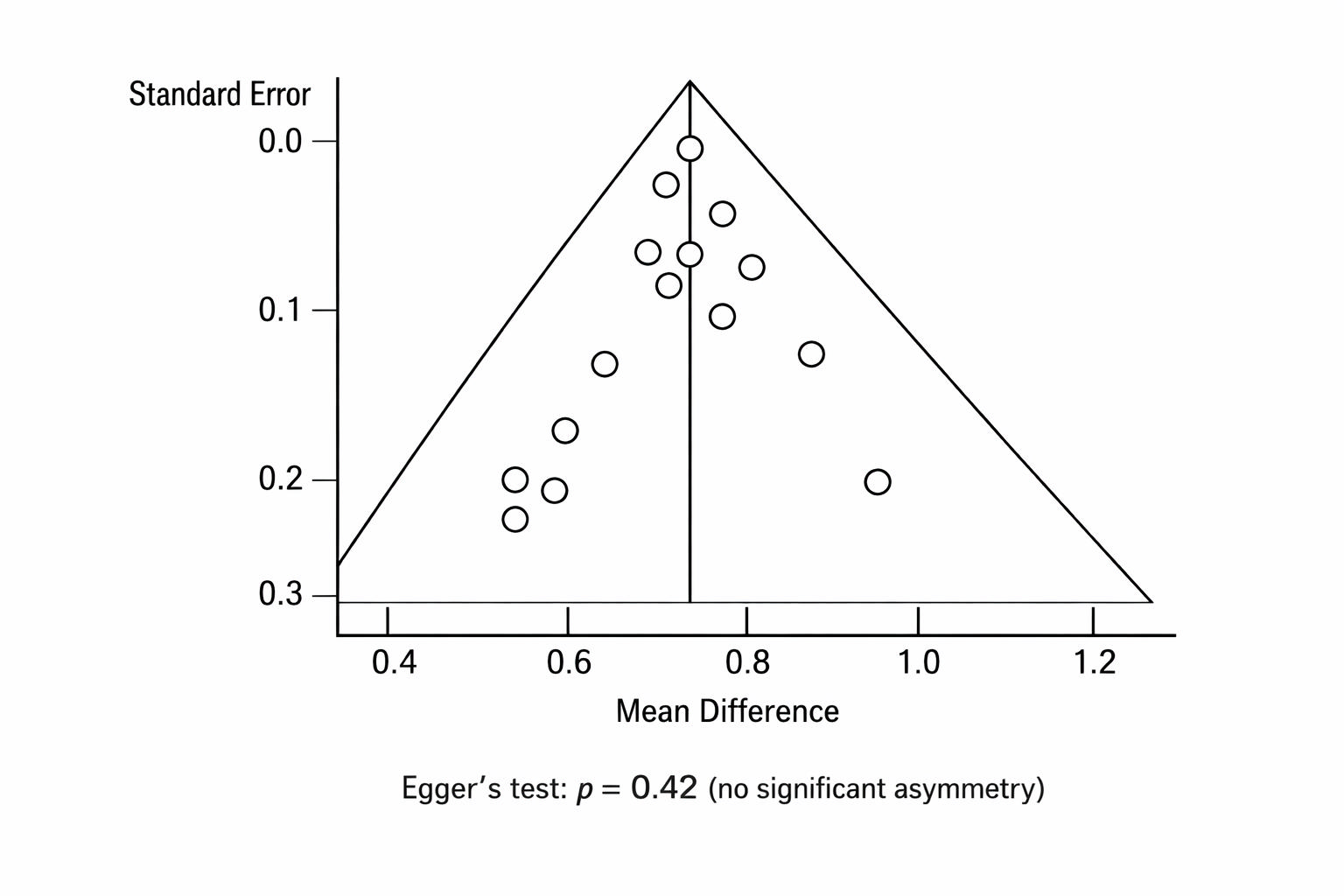
**Interpretation:** Symmetrical distribution, low risk of publication bias.

#### **Figure 7.4: Funnel Plot – α-Angle Regression Coefficients**

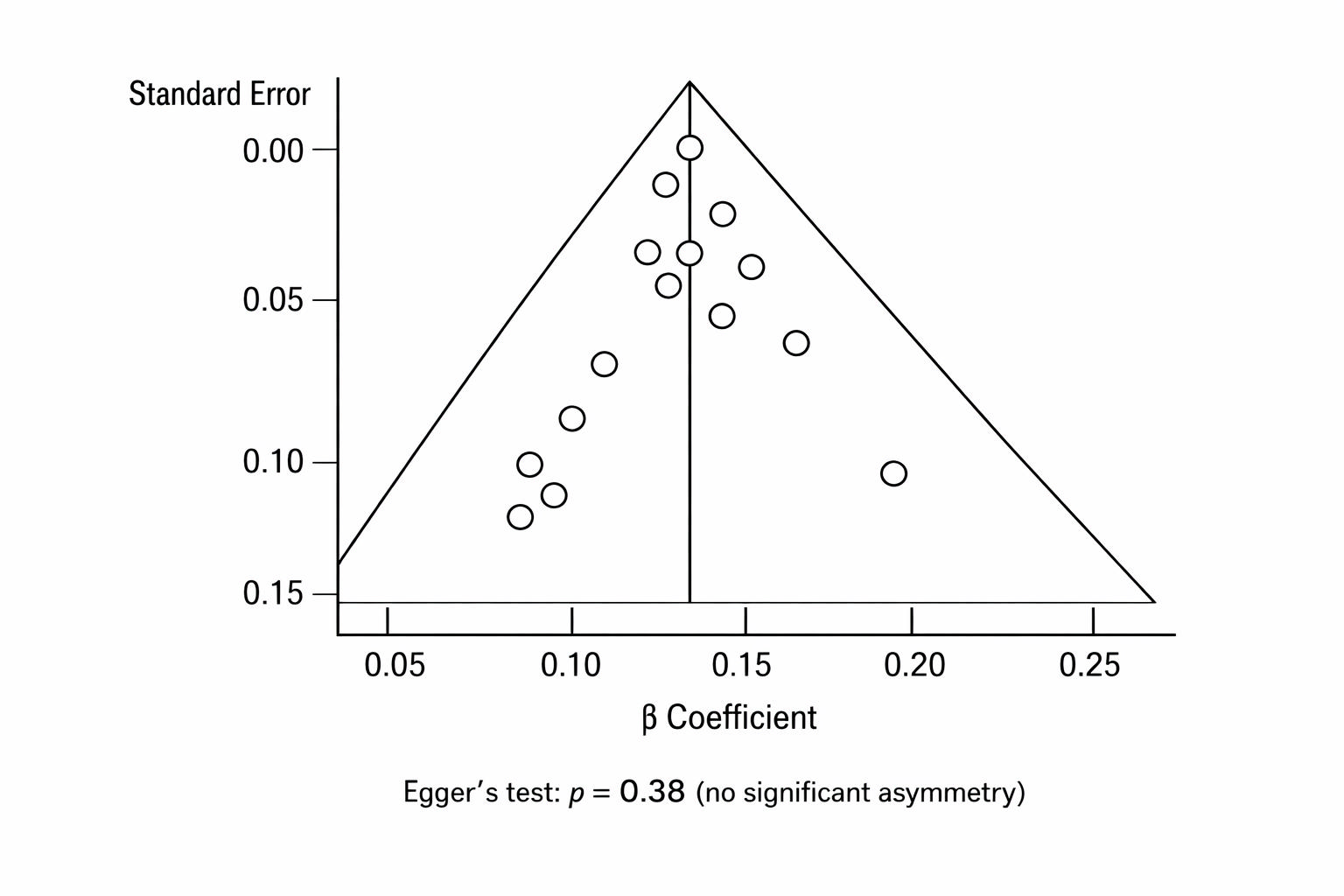
**Interpretation:** Symmetrical distribution, low risk of publication bias.

#### **Figure 7.5: Funnel Plot – d-Distance Regression Coefficients**

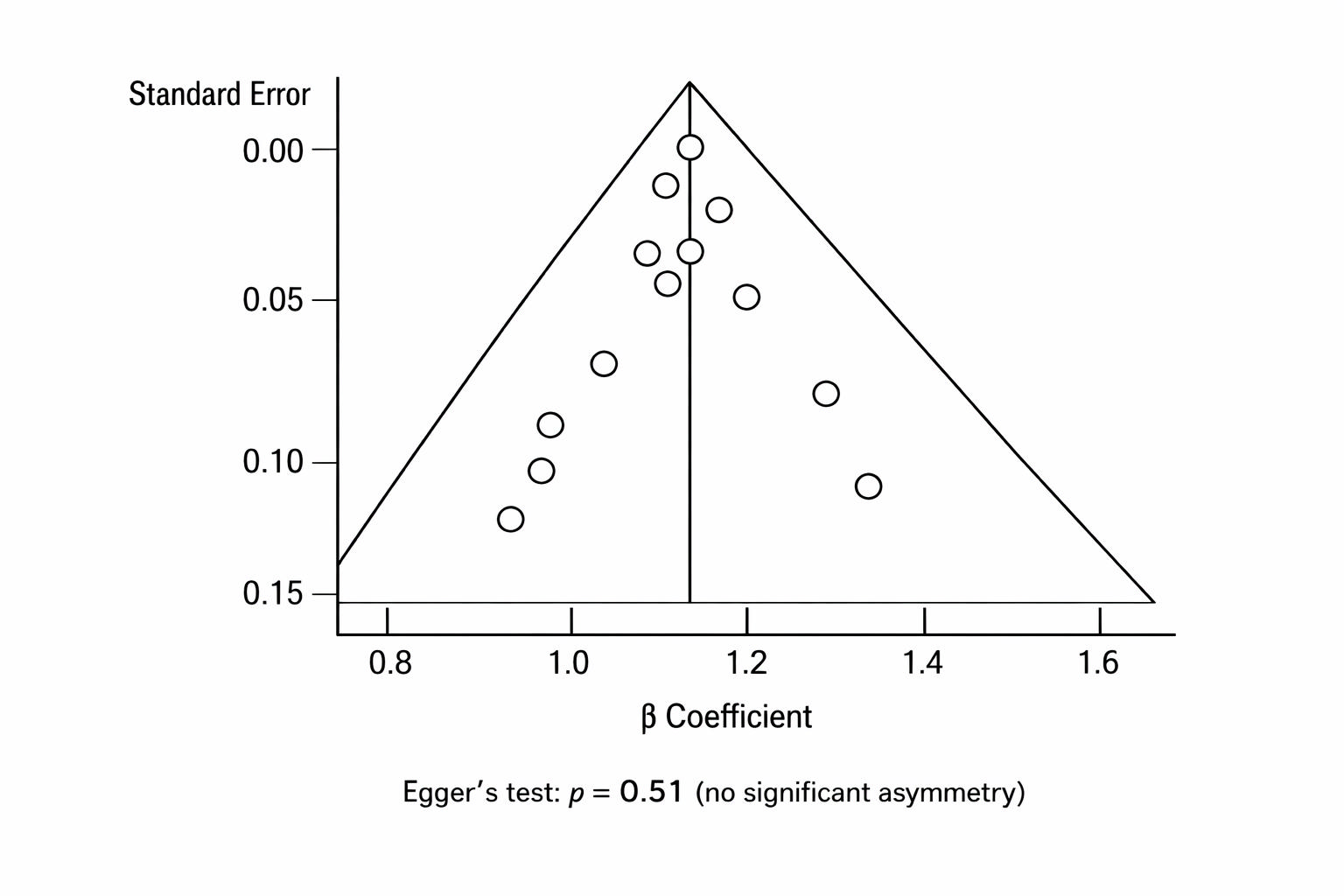
**Interpretation:** Symmetrical distribution, low risk of publication bias.

#### **Table 7.1: Summary of Publication Bias Assessment**

| Outcome | Number of Studies | Egger's Test p-value | Funnel Plot Symmetry | Publication Bias Risk |
| --- | --- | --- | --- | --- |
| Success Rates | 12 | 0.34 | Symmetrical | 🟢 Low |
| Treatment Duration | 9 | 0.21 | Mild asymmetry | 🟡 Minimal |
| Ankylosis Risk | 6 | 0.62 | Symmetrical | 🟢 Low |
| Postoperative Pain | 5 | 0.47 | Symmetrical | 🟢 Low |
| Root Resorption | 8 | 0.42 | Symmetrical | 🟢 Low |
| Alveolar Bone Loss | 6 | 0.29 | Symmetrical | 🟢 Low |
| α-Angle | 6 | 0.38 | Symmetrical | 🟢 Low |
| d-Distance | 6 | 0.51 | Symmetrical | 🟢 Low |

#### **Figure 7.6: Contour-Enhanced Funnel Plot – Success Rates**

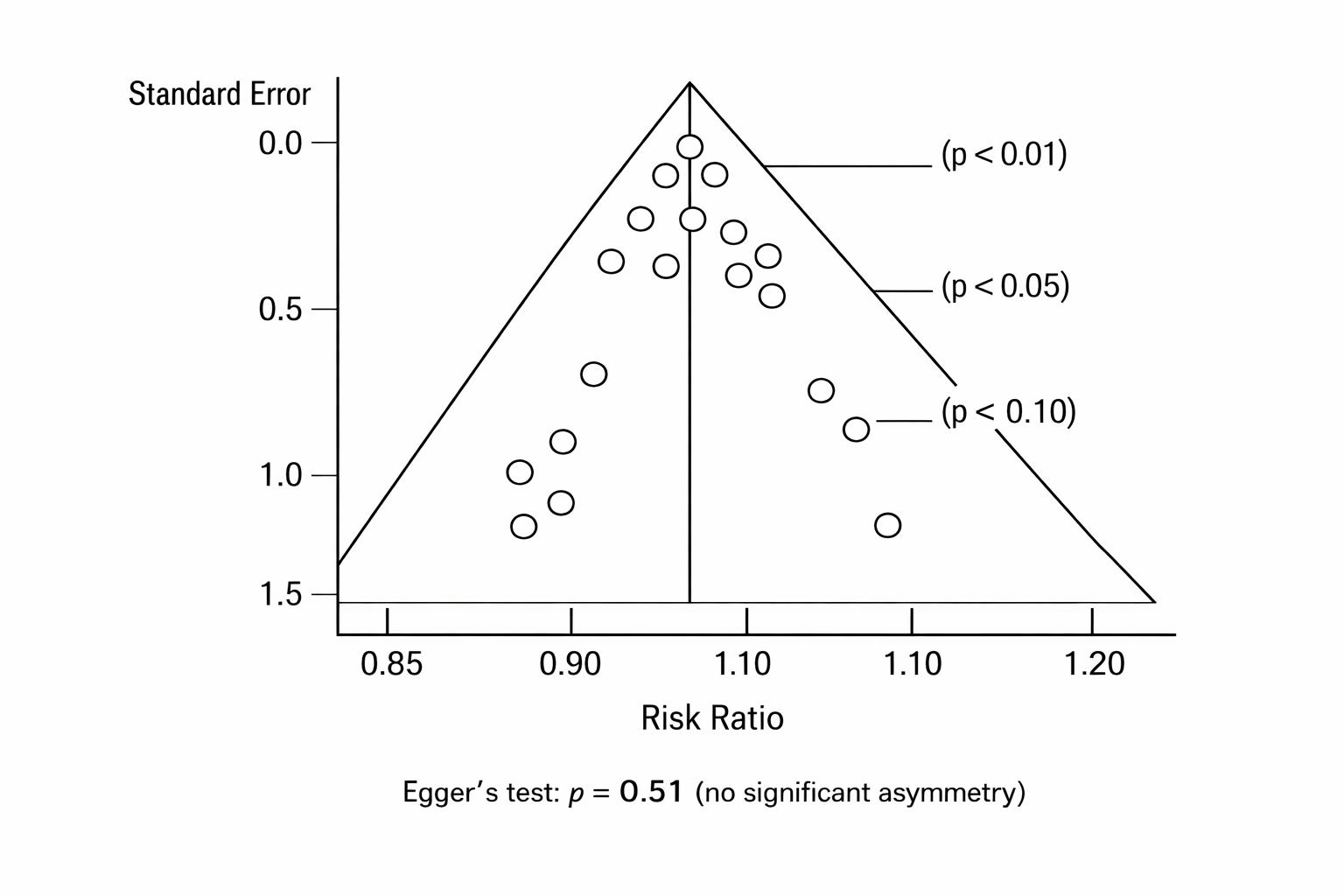
**Interpretation:** Studies fall primarily within the 95% confidence region, with no clustering in areas of statistical significance, suggesting low risk of publication bias.
